# An ultrasensitive method for detecting mutations from short and rare cell-free DNA

**DOI:** 10.1101/2023.03.14.23287139

**Authors:** Lin Wang, Yu Zhuang, Yue Yu, Zhiwei Guo, Qiaomei Guo, Lihua Qiao, Xueqing Wang, Xiaohui Liang, Pengpeng Zhang, Qifan Li, Chenjun Huang, Rong Cong, Yinghui Li, Bin Che, Guomin Lin, Mingming Rao, Rongjun Hu, Jiatao Lou, Wei Wang, Guohua Yang

## Abstract

**Background:** Cell-free DNA (cfDNA) promises to serve as surrogate biomarkers for non-invasive molecular diagnostics. Disease-specific cfDNA, such as circulating tumor DNA (ctDNA), was short and rare, making the detection performance of the current targeted sequencing methods unsatisfying.

**Methods:** Through introducing a linear pre-amplification process and optimizing the adapter ligation with customized reagents, we developed the One-PrimER Amplification (OPERA) system. In this study, we examined its performance in detecting mutations of low variant allelic frequency (VAF) in various samples with short-sized DNA fragments.

**Results:** In cell line-derived samples containing sonication-sheared DNA fragments with 50-150 bp (peak at 70-80 bp), OPERA was capable of detecting mutations as low as 0.0025% VAF, while CAPP-Seq only detected mutations of >0.03% VAF. Both single nucleotide variant and insertion/deletion can be detected by OPERA. In synthetic fragments as short as 80 bp with low VAF (0.03%-0.1%), the detection sensitivity of OPERA was significantly higher compared to that of droplet digital polymerase chain reaction. The error rate was 5.9×10^−5^ errors per base after de-duplication in plasma samples collected from healthy volunteers. By suppressing “single-strand errors”, the error rate can be further lowered by >5 folds in *EGFR* T790M hotspot. In plasma samples collected from lung cancer patients, OPERA detected mutations in 57.1% stage I patients with 100% specificity and achieved a sensitivity of 30.0% in patients with tumor volume of less than 1 cm^3^.

**Conclusions:** OPERA can effectively detect mutations in rare and highly-fragmented DNA.

**Trial registration:** This study has been registered on ChiCTR (ChiCTR1900024028) at 23^rd^ June 2019. Keywords: cell-free DNA; library preparation; liquid biopsy; mutation; next-generation sequencing.

## Background

Tremendous efforts have been made in the past decade to improve the sensitivity, accuracy and throughput of molecular diagnostics [1]. Cell-free DNA (cfDNA) has been suggested to be a promising surrogate biomarker for non-invasive molecular diagnostics. Recent findings demonstrated that the cfDNA with a short size was highly associated with diseases [2]. Due to the action of nucleases, circulating cfDNA is fragmented [3,4], especially for abnormal DNAs such as circulating tumor DNA (ctDNA), pathogen-derived DNA and fetal DNA, those abnormal DNAs are quite rare and their fragment lengths are particularly short [3,5-8]. Loss of rare and short-sized cfDNA could lead to a lower sensitivity for the detection of disease-associated cfDNA. Moreover, these target DNAs in liquid sample can be masked by a large amount of background DNA from hematopoietic cells, making detection of these target DNA in cfDNA even more challenging.

The efficiency of cfDNA detection by targeted sequencing methods could be affected by two important factors. The first factor is “theoretical detection ratio”, which represents the proportion in theory for randomly fragmented molecules to be detected. As calculated in Fig. S1, the ratio is related to the length of the detected DNA molecule (Fragment length) and the length of the sequence required for detection (Target length) [9]. For PCR-based technologies, the shortest length of the specific target region is around 60bp (forward and reverse primers +probe, 20 bp each); while the probe length in hybrid capture technology is mostly about 120bp (e.g. CAPP-Seq). Therefore, the theoretical detection ratio for these methods is low for short DNAs. The second factor associated with the cfDNA detection efficiency is the “library conversion rate” which is related to the ligation efficiency during library construction, the molecular loss during the purification after ligation and the amplification effect before ligation. For most hybrid capture technologies that are not amplified before ligation, the library conversion rate is generally not higher than 30%, which accounts for the main limiting factor [10]. The final detection efficiency is approximately the product of “theoretical detection ratio” and “library conversion rate”.

PCR-based and hybrid capture-based methods are nowadays the most common principle applied in nucleic acid-based molecular technologies [1], but due to the low “theoretical detection ratio” and “library conversion rate” [11-13], neither of them can circumvent the problem of short and rare DNAs. In this study, we developed a tailor-made system for detecting rare, fragmented DNA, named One-PrimER Amplification (OPERA), and evaluated its performance in identifying mutations including single nucleotide variant (SNV) and insertion/deletion (Indel).

## Methods

### Blood and saliva samples from healthy volunteers

Peripheral blood samples and unstimulated saliva samples were collected from healthy volunteers (without a prior history of cancer) using EDTA-containing vacuum tubes (BD Biosciences) and 5 mL centrifuge tubes (Eppendorf), respectively. Peripheral blood samples were temporarily stored at 4 °C and processed within 24 hours after collection. Whole blood samples were first centrifuged at 1200 g for 10 min. Then, the supernatant was transferred to a new 5 mL centrifuge tube and subsequently centrifuged at 13000 g for 10 min. The supernatant, which was the plasma sample, was again transferred to a new EP tube and stored at -80 °C before analysis. Plasma cfDNA was extracted by QIAamp® Circulating Nucleic Acid Kit (Qiagen, Germany) according to the instruction manual. The cell pellet after the first centrifugation was also collected as blood cell samples. These blood cell samples were stored at -80 °C before analysis. Blood cell and saliva genomic DNA (gDNA) was extracted by HighPure PCR Template Preparation kit (Roche Diagnostics) according to the instruction manual.

To mimic the plasma cfDNA samples, sonication-sheared (Covaris, S220, USA) gDNA (roughly 150bp) derived from the blood cells of healthy volunteers after quantification with Qubit (Thermo Fisher Scientific) was used as control DNA fragments.

### Cell line-derived samples

DNA standards for mutation detection were extracted from cancer cell lines. Sonication was also performed in DNA standards, resulting in two types of DNA fragment lengths: short fragments with DNA lengths of 50-150 bp (peak at 70-80 bp) and long fragments with DNA lengths of 150-300 bp (peak at 180 bp). DNA fragments were prepared by agarose gel electrophoresis. Multiplex DNA standard covers 10 types of hotspot mutations: *BRAF* V600E (purchased from Cobioer, GBD33766601), *EGFR* delE746-750 (exon 19 deletion; 19del; from HCC827 cells), *EGFR* T790M (from H1975 cells), *EGFR* L858R (from H1975 cells), *ERBB2* insA775-G776YVMA (exon 20 insersion; 20ins; purchased from Cobioer, GBD43448227), *KRAS* Q61H (purchased from Cobioer, CGD11749985), *KRAS* G12C (from H23 cells), *NRAS* Q61H (purchased from Cobioer, CGD11749985), *PIK3CA* E545K (purchased from Cobioer, CGD23175411) and *TP53* S215G (from HCC827 cells). To prepare reference samples with VAF of 0.0025%, 0.05%, 0.01%, 0.03%, 0.1%, 0.3% and 1%, the standard DNA (VAF=1%) was spiked into control DNA fragments. The control DNA fragments itself (not sheared by sonication) served as control in determination of somatic mutations. 30-40 ng of DNA was used for each test, with 4 replicates.

### Synthetic DNA fragments

Synthetic DNA fragments of *EGFR* L858R or T790M mutations with lengths of 80, 120 and 160 bp were synthesized by Shanghai Bioligo Biotechnology Co., Ltd. (Shanghai, China). As shown in Fig. S2, all the synthetic fragments contain a core 13-17 bp sequence. To mimic the randomly digested DNA fragments present in blood specimens, the synthetic fragments with a unique length are regularly spaced by 6-13 bp, equimolarly mixed based on OD260 quantification (Nanodrop, Thermo Fisher Scientific) and then spiked separately with different input into plasma cfDNA samples obtained from healthy volunteers, resulting in samples of 1%, 0.3%, 0.1%, 0.03% and 0% variant allelic frequencies (VAF). 20-40 ng of DNA was used for each test, with 4 replicates.

### Clinical specimens from lung cancer patients

Matched tumor tissue specimens and whole blood samples were collected from lung cancer patients recruited from Jiangsu Province Hospital under a prospective clinical trial (Registration number: ChiCTR1900024028). The inclusion criteria were as follows: 1) Aged between 18 and 80 years old; 2) Patients with suspicious non-small cell lung cancer (NSCLC) and planned to receive surgical treatment; 3) Confirmed NSCLC patients or benign lung disease patients; 4) Stage I-IIIA (for NSCLC patients); 5) Sufficient amount of samples available for analysis; 6) Signed informed consent obtained. The exclusion criteria were as follows: 1) Recurrent NSCLC; 2) Lysis of blood samples; 3) Blood samples were not processed into plasma and blood cell samples within 24 hours; 4) Pregnant or lactating patients; 5) Received any anti-tumor therapy before surgery; 6) Patients with a prior cancer history. The clinical trial was conducted in accordance with the Declaration of Helsinki.

### Processing of clinical specimens

In detail, tumor tissue specimens were collected during surgery resection. Tumor tissue was dissected into appropriate size (half of the size of a soybean) and stored in a cryogenic vial (Corning, Germany) temporarily at 4 °C. Pathology assessment based on hematoxylin-eosin staining was performed by experienced pathologists to confirm the proportion of cancer cells present in the tumor tissue samples. All the tumor tissue samples had a cancer cell proportion of over 80%. Additionally, adjacent normal tissue samples without any cancer cells were dissected into appropriate size and stored in a cryogenic vial by the pathologist. Both the tumor tissue and adjacent normal tissue samples were stored at -80 °C before analysis. gDNA of these samples were extracted by using HighPure PCR Template Preparation kit (Roche, Switzerland) according to the instruction manual. Ten milliliter peripheral blood samples were collected before surgery. Plasma cfDNA and blood cell gDNA samples were prepared from the blood samples using the same procedure as described in the preparation of samples of healthy volunteers.

The formalin-fixed paraffin-embedded (FFPE)-processed adjacent normal tissue specimens for testing of the paired primer system were also collected from the same group of lung cancer patients recruited from Jiangsu Province Hospital. All the samples were examined by hematoxylin-eosin staining and observed by experienced pathologists once again to further confirm that no cancer cells are present. gDNA of these samples were extracted by using QIAamp DNA FFPE Tissue Kit (Qiagen) according to the instruction manual. Before detection, all gDNA samples were randomly fragmented by DNase I using a DNA fragmentation kit (Apogenomics, Shanghai, China) according to the instruction manual.

### CAPP-Seq

CAPP-Seq was performed as previously described [12]. The targeted regions (approximately 204 kbp) for CAPP-Seq were listed in Table S1. The sequencing depth was 20,000-25,000x and the average read depth after de-duplication was 2,000x.

### Droplet digital PCR

The *EGFR* L858R and T790M mutation detection assay for droplet digital PCR (ddPCR) were purchased from Bio-Rad (PrimePCR ddPCR Mutation Detection Kit, Catalog no.: 1863104 & 1863103). ddPCR was performed using QX200 Droplet Digital^™^ PCR system (Bio-Rad, Hercules, USA) according to the instruction manual. A 20 μL reaction mixture was prepared comprising of 10 μL ddPCR Supermix^™^ for probes (Bio-Rad), 8 μL primers and probe mix, and 2 μL DNA. The final concentration of primers and probe was 600 nM and 300 nM, respectively. The amplification conditions were 10 min DNA polymerase activation at 95 °C, followed by 40 cycles of a two-step thermal profile of 30 seconds at 94 °C for denaturation, and 60 seconds at 64 °C for annealing and extension, followed by a final hold of 10 min at 98 °C for droplet stabilization, and cooling to 4 °C. The temperature ramp rate was set to 2.0 °C/s, with the lid heated to 105 °C, according to the Bio-Rad recommendations. The cutoff value of a positive call for ddPCR was defined as the highest detected value (in integral number) in negative control samples for each variant. For example, when the highest detected value in negative control samples was 1.8, the cutoff value was set as 2; when the highest detected value in negative control samples was 2.0, the cutoff value was set as 3.

### Procedures for OPERA

Briefly, the procedures for OPERA can be divided into five key steps: 1) linear amplification, 2) adapter ligation, 3) indexing PCR, 4) PCR amplification, and 5) sequencing. Majority of molecular biology reagents were contained in the OPERA Jupiter® library construction kit (Apogenomics). Primers and adapters were synthesized by Shanghai Bioligo Biotechnology Co., Ltd, and the sequences were shown in Table S2.

In linear amplification, fragmented DNA samples (30-40 ng) were mixed with 1 μL of 20× dualistic primer (DL primer) mixture, 4 μL of 5× APO Buffer I, 0.8 μL of APO biotin-dNTP mix 1 and 0.2 μL of APO-Enchanted^™^ DNA polymerase I. The DL primers comprise a SP1 sequence and a targeting sequence that is complementary to the target regions (Table S2). For all the DL primers, the –OH group of 3’ terminal nucleotide is substituted by a DL blocker, which is a nucleotide analogue with 3’ C3 Spacer modification. In addition, the last three nucleotides at the 3’-end of the DL primer possess phosphorothioate modifications. APO-Enchanted DNA polymerase I is a high-fidelity DNA polymerase that is able to cleave the DL blocker with its 3’-5’ exonuclease activity and to incorporate biotinylated dNTPs. The reaction mixture underwent amplification using the Applied Biosystem Veriti 96-well thermal cycler (ThermoFisher Scientific). The reaction condition was as follows: 1) Pre-denaturation at 98 °C for 100 seconds; 2) 30-35 cycles of denaturation at 95 °C for 10 seconds, annealing at 67 °C for 15 seconds, extension at 70 °C for 60 seconds. The single-stranded DNA (ssDNA) product was purified by Dynabeads MyOne Streptavidin C1 (ThermoFisher Scientific) according to the instruction manual. The number of cycles of linear amplification is dependent on the sample type. Thirty cycles of linear amplification is preferred for gDNA fragment and synthetic DNA samples, while 35 cycles is preferred for cfDNA samples.

In adapter ligation, 20 μL of purified ssDNA products were mixed with 1.25 μL of single-stranded adapter, 2.5 μL of 10× ligase buffer, 0.63 μL of MnCl_2_, and 0.63 μL of ssDNA ligase. The single-stranded adapters comprise a unique molecular identifier (UMI) sequence and a sample barcode sequence (index i7), and thus should be added independently in each experiment (Table S2). The 5’ terminus of the adapter is phosphorylated and the 3’ terminus of it contains a C3-Spacer group substitution. The ligation reaction on PCR thermal cycler was as follow: 1) 60 °C for 60 min; 2) 90 °C for 3 min.

In indexing PCR step, 24 μL of ligated products were mixed with 1 μL of forward primer (i5 index primer; 100 nM), 1 μL of reverse primer (P7 primer; 200 nM), 10 μL of 5× SLA Buffer, 1 μL of dNTP Mix (200 μM), 0.25 μL of SLA^™^ high-fidelity DNA polymerase, and 12.75 μL of ddH2O. The forward primer contains a sample barcode sequence (index i5) and thus should be added independently in each experiment. The PCR reaction condition was as follows: 1) Pre-denaturation at 95 °C for 3 min; 2) 5 cycles of denaturation at 95 °C for 10 seconds, annealing at 70 °C for 10 seconds, extension at 72 °C for 30 seconds; 3) extension at 72 °C for 120 seconds and temporarily stored at 8 °C. The pre-amplified product was purified by AMPure XP magnetic beads (Beckman Coulter) according to the instruction manual.

In PCR amplification, 20 μL of pre-amplified products were mixed with 0.6 μL of forward and reverse primers (p5 or p7 primer; 10 μM each), 6 μL of 5× SLA Buffer, 0.6 μL of dNTP Mix (10 mM each), 0.15 μL of SLA TM high-fidelity DNA polymerase, and 2 μL of water. The reaction condition was as follow: 1) Pre-denaturation at 95 °C for 3 min; 2) 15 cycles of denaturation at 95 °C for 10 seconds, annealing at 72 °C for 10 seconds, extension at 72 °C for 30 seconds; 3) extension at 72 °C for 120 seconds and temporarily stored at 8 °C. The library molecules were purified by AMPure XP magnetic beads according to the instruction manual. The purified library samples were quantified by OPERA^™^ library quantification kit (Apogenomics) and mixed for sequencing.

In sequencing, prepared libraries underwent 150 bp paired-end sequencing (100,000x) using the Illumina NovaSeq 6000 service platform (Mingma Technologies, Shanghai, China).

### Panels and primer design

The criteria for designing the DL primers were as follows: 1) the primer length should be ranged from 20 nt to 40 nt; 2) the distance between the 3’-end of the primer and the target site/region should be as close as possible and ranged from 5-55 bp; 3) access the specificity of the primers with Blast; 4) the melting temperature of each primer should be >64 °C in the condition of [Na+] = 100 mM, [Mg2+] = 0 mM, and [primer] = 100 nM; 5) the homodimer and heterodimer probability of all primers in each panel should be analyzed, a primer pair with a strong delta G (< -9 kcal/mol) should be avoided.

The detailed primer sequences of each panel are shown in Table S3.

### Bioinformatic analysis of OPERA sequencing data

Paired-end sequencing reads were first trimmed using Cutadapt version 1.18 to remove adapters, and then aligned to human genome HG38 using BWA mem version 0.7.13, and sorted with SAMtools. If the R1 read of one read pair was mapped from the start position to 5 bp downstream of a target-specific DL primer, then the read pair was considered to be on-target. Both the positions of R1 and R2 reads were used to observe the length of the template DNA. The reads from the same original molecule were identified with a de-duplication strategy, which grouped the reads according to the UMI sequence and the positions of R1 and R2 reads, and called the molecular consensus sequences from paired-end reads with the same unique molecular tag requiring minimum two reads to support a consensus base. Fragment length, sequencing depth and UMI counts were calculated using a custom script developed in Python 3.7.1 and Perl 5.16.3. To detect SNVs and Indels, VarScan 2 was applied with the options --min-reads22 and --min-avg-qual 30 to improve the confidence of variant call.

### Calculation of background error rates

The background error rates were measured by sequencing the paired plasma and saliva samples from healthy volunteers and the FFPE-processed adjacent normal tissue specimens as previously mentioned. The 50 bp downstream of a target-specific DL primer of the on-target reads were evaluated with the mutation panel. For the single-strand error rates, the overlap of the sequencing region by paired primers in two different paired primer detection system respectively was used for calculation of error rates. We excluded the positions in which the count of one base is only one and the reads whose final base possessed mismatch. Non-reference bases whose allele frequency is >1% were also excluded. Once the positions and alleles met the above criteria, the background error rate was defined as the fraction of non-reference bases in all sequenced bases.

### Statistical analysis

The significant difference of error rate which detected by the plus panel and minus panel was examined by paired student t test. The correlation between plasma VAF and the tumor volume or the product of tumor volume and tumor tissue VAF is tested by nonparametric Spearman correlation analysis. A two-sided *P* value of less than 0.05 was considered as statistically significant.

## Results

### Principle of the OPERA system

The detailed procedures of OPERA are shown in Fig. 1. To maximize “library conversion rate”, a linear amplification before ligation was performed. Furthermore, by using single primer to amplify short target DNA with random breakpoint, OPERA system minimized the target length to be as short as 42 bp (55 bp in average), leading to the theoretically highest “theoretical detection ratio”. To prevent the ligation between the free primer and adapter, we developed the DL primer and APO-enchant polymerase. The DL primer comprises a DL Blocker at its 3’-end, and exists in the form of two different states: the bound DL primer and the free DL primer, as shown in step 1 and 2 of Fig. 1 respectively. When the DL primer is free from the template DNA, the DL Blocker prevents its ligation with the adapter in the next step. Once the DL primer binds to a target DNA, the DL blocker can be cleaved by the APO-Enchant polymerase which possesses 3’-5’ exonuclease activity, then the DL primer is activated to initiate the extension with the target DNA as template. Since the ligation only occurs in 3’ terminal of primer extension product and 5’ terminal of adapter, this effectively avoids the originally dominant by-products caused by ligation of free primers and adapters. Together, these features allow reliable and ultrasensitive detection of rare and short DNA molecules. After ligation, the single strand library molecule was amplified with indexed primers, and finally formed a conventional double strand library molecule (Fig. 1), which is suitable for Illumina next-generation sequencing (NGS) platform [14]. In sonication-sheared DNA samples derived from blood cells of healthy volunteers, we observed a 25-fold increase in ligation efficiency and a 22-fold increase in library conversion rate for DL primer compared to primer without modifications (Table S4), proving that the modifications of DL primer can effectively improve OPERA’s detection performance.

**Fig. 1.**
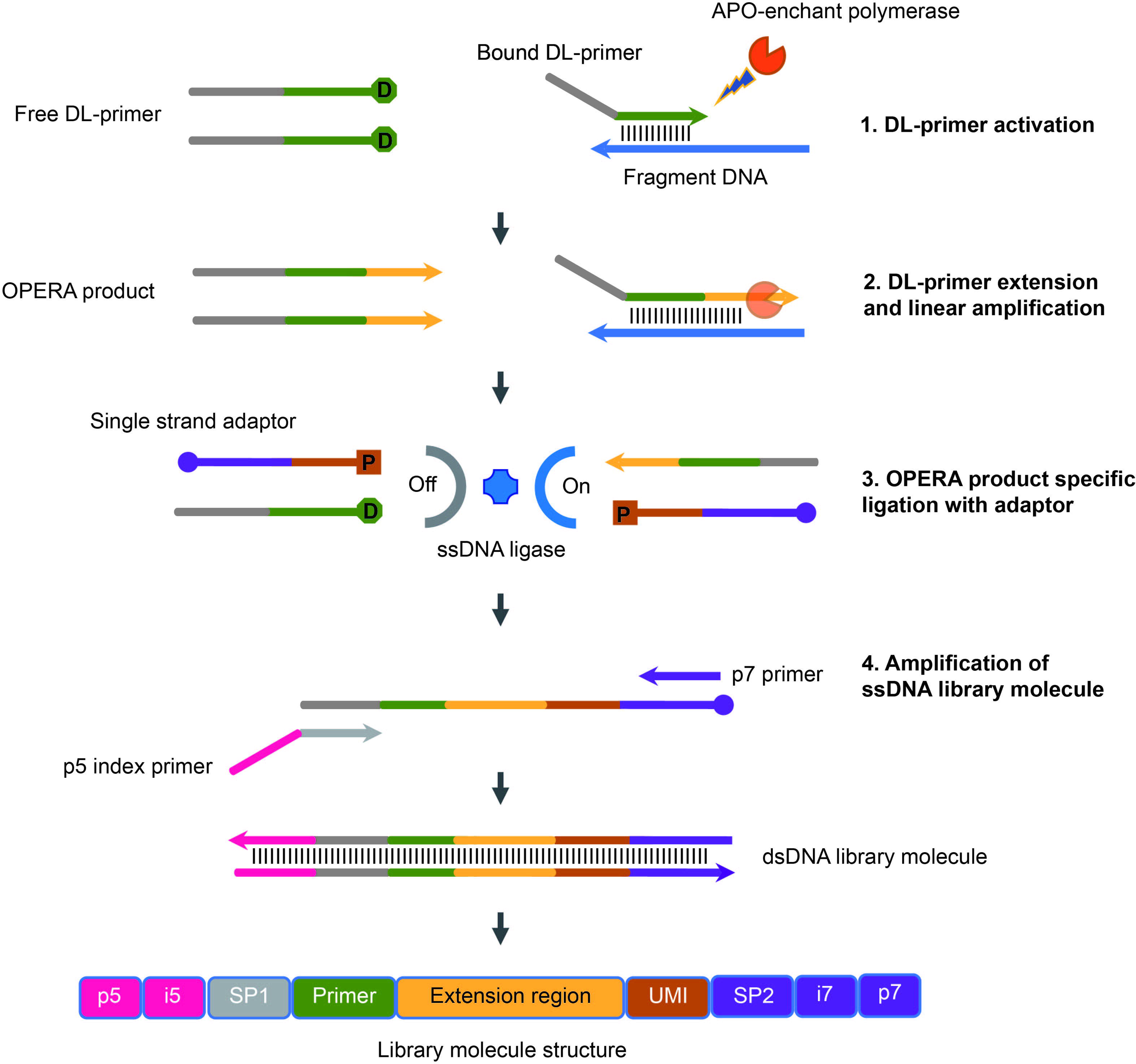
Principle of the OPERA system. Schematic diagram of the OPERA workflow. “D” in the green circle represents the DL blocker at the 3’-end of the DL primer, “P” in the red square at the 5’-end and the purple circle at the 3’-end of the single-strand adapter represent a phosphate group and a blocker, respectively. UMI: Unique Molecular Identifier. SP1: Sequencing Primer for Read 1. SP2: Sequencing Primer for Read 2. SP1, SP2, p5, p7, i5 and i7 are compatible with Illumina NGS platform.

### Performance of OPERA in short and rare fragment detection

To explore the detection sensitivity of OPERA, mutated DNA samples derived from cancer cell lines were spiked into control cfDNA samples at different ratios. CAPP-Seq as one of the most widely applied hybrid capture-based sequencing methods was used as a comparator. For short DNA fragments (50-150 bp), the detection sensitivity was generally higher in OPERA compared to CAPP-Seq at mutation level (Fig. 2A). OPERA was capable of detecting mutations as low as 0.0025% VAF, while CAPP-Seq only detected mutations of >0.03% VAF. Both SNVs and Indels can be effectively detected by OPERA (Table S5). For long DNA fragments (150-300 bp), the detection sensitivity was still higher in low VAF (0.0025%-0.1%), but comparable in high VAF (0.3%-1%; Fig. 2B). Additionally, the mean VAF detected by OPERA was strongly correlated with the expected VAF of the samples (Fig. S3), suggesting that OPERA gives quantitative results.

**Fig. 2.**
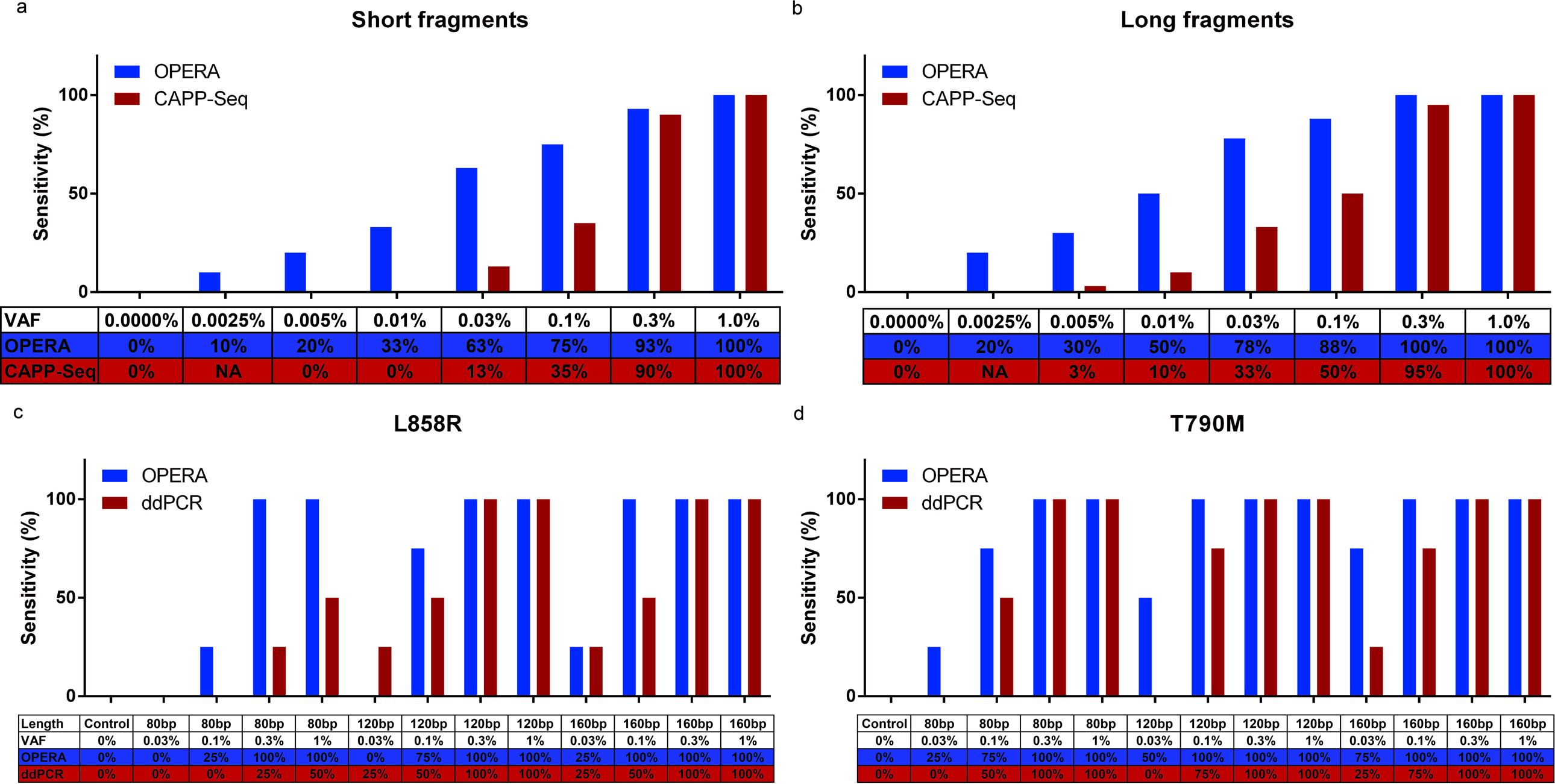
Performance of OPERA in cell line-derived and synthetic DNA fragments. Comparison of the detection sensitivity of OPERA and CAPP-Seq in cell line-derived DNA fragments with different VAFs: (a) short DNA fragments with 50-150 bp; (b) long DNA fragments with 150-300 bp. Comparison of the detection sensitivity of OPERA and ddPCR in synthetic DNA fragments containing (c) *EGFR* L858R and (d) *EGFR* T790M mutations, with different fragment lengths and different VAFs. Each experiment was repeated 4 times. The error bars indicate standard deviations.

To further examine the ability to detect random short fragments, two types of synthetic DNA (containing *EGFR* L858R and T790M mutations, respectively) with lengths of 80, 120 and 160 bp were spiked separately in plasma cfDNA samples obtained from healthy volunteers and tested by OPERA. ddPCR served as a comparator. To maximize the ability to detect short fragments, specific primers were designed to be as short as possible: the DL primer for OPERA was 30 nt (L858R) and 28 nt (T790M) in length. In samples of 80 bp-fragment and low VAF (0.03%-0.1%), the detection sensitivity of OPERA was significantly higher compared to that of ddPCR (Fig. 2C, 2D and Table S6), indicating that OPERA could better detect mutations present in short DNA fragments compared to ddPCR.

### Fidelity of OPERA

To verify the fidelity of OPERA, plasma cfDNA samples from 20 healthy volunteers were tested with a panel designed to detect hotspot mutations in lung cancers. The paired saliva samples were used as a control group to evaluate the effect of clonal hematopoiesis in plasma cfDNA. The error rate of OPERA was 5.9×10^−5^ and 7.9×10^−5^ errors per base after de-duplication with UMI in plasma and saliva, respectively, which were 24 to 32 folds lower than that before de-duplication (Fig. 3A). It was found that C>T was the major type of error for the actual detected strand (Fig. 3B and C). These observations were in agreement with a previous study using strand specific error analysis [13].

**Fig. 3.**
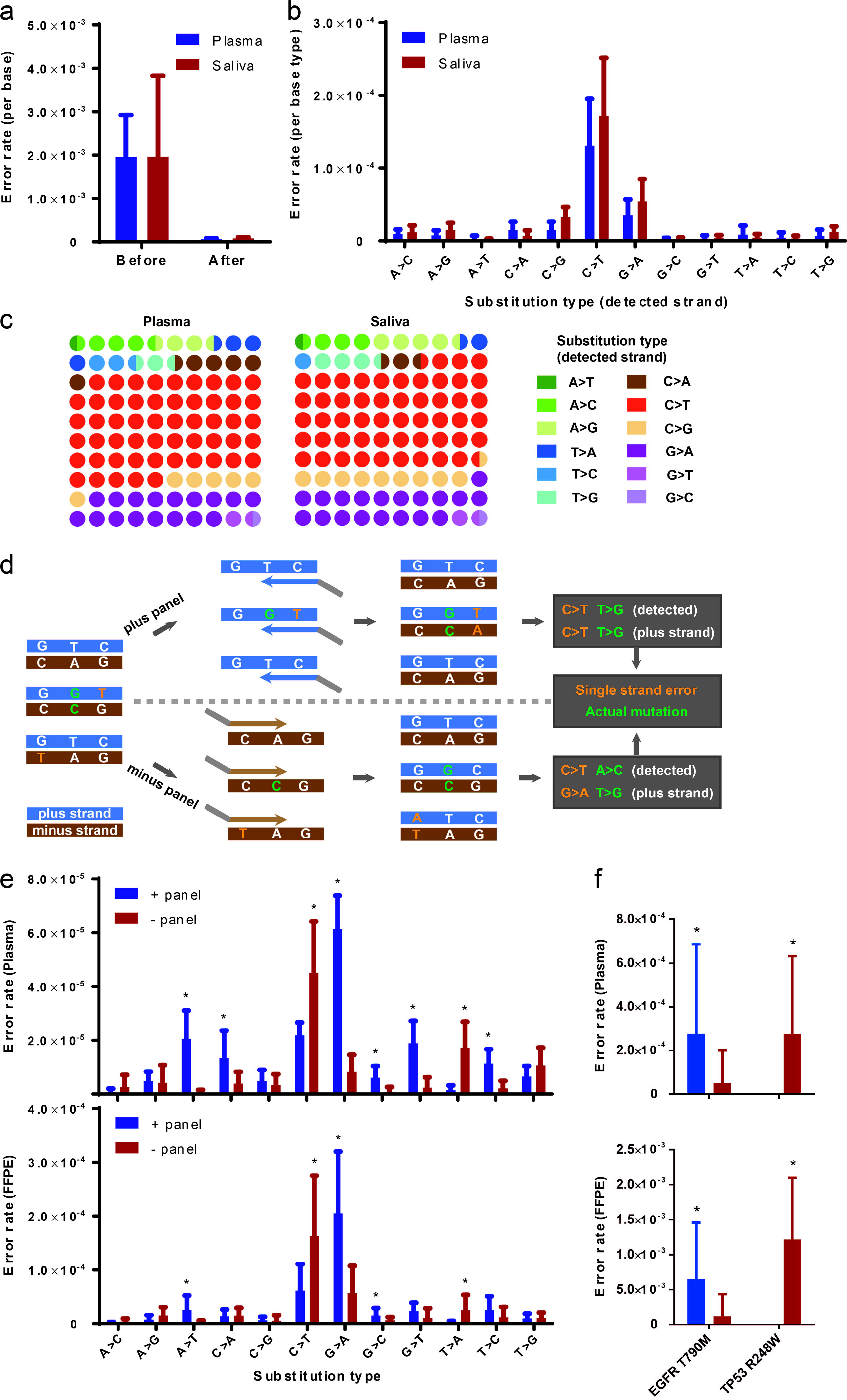
Distinction of single-strand errors by OPERA. (a) Comparison of the error rate (error per base) before and after de-duplication. (b-c) Types of errors. Bar chart and proportional plot showing the types of errors in terms of (b) error per base and (c) error count, respectively. Errors are distinguished based on the reference sequence of actual detected strand. (d) Schematic representation of errors based on the different reference sequences (detected strand and plus strand). The “plus panel” or “minus panel” indicate the panel that bound to plus strand or minus strand, respectively. Blue strand represents the “+” strand, and brown strand indicates the “-” strand. The base in green represents an actual mutation that occurs in both strands, while the base in yellow shows the single-strand error. (e-f) Types of error rate of (e) all region and (f) representative hotspots (*EGFR*-T790M and *TP53*-R248W) as determined by the + panel (bound to plus strand) and the -panel (bound to minus strand) in blood ctDNA and FFPE-processed adjacent normal tissue samples, respectively. Errors are distinguished based on the reference sequence of plus strand. The significant difference of error rate detected by the 2 strand-specific panels was examined by paired t test. Asterisk represents *P* value of less than 0.05.

As a single primer amplification system, OPERA is capable of designing strand-specific primers (targeting either the plus or minus strand) to avoid single-strand originating errors. Therefore, we developed a paired strand-specific primer system. Eleven pairs of primers targeting the same region were divided into two strand-specific panels, which were used to detect hotspot mutations that are possibly affected by single-strand errors (i.e. *EGFR* T790M) (Fig. 3D). Blood cfDNA samples from healthy volunteers and FFPE-processed adjacent normal tissue specimens were used to evaluate the single-strand error by the paired primer system (results of blood cell and saliva samples were shown in Fig. S4). We found that, no matter in blood cfDNA or FFPE samples, there exists a significant strand-bias error rate for the majority of substitution types, especially for C>T and G>A errors (Fig. 3E). Using the plus strand sequence as reference, substitution type C>T preferentially biased toward the “+” strand while G>A errors biased toward the “-” strand. These results were actually in agreement with data of Fig. 3B, in which the detected strand sequence was used as reference. These suggested that, through targeting the specific strand, OPERA can dramatically reduce these single-strand errors based on specific substitution types. For instance, when the appropriate strand-specific primer panel was used, the error rate can be lowered by >5 folds in *EGFR* T790M, and that of *TP53* R248W even reduced to 0, in blood cfDNA and FFPE samples (Fig. 3F; results of blood cell and saliva samples were shown in Fig. S4).

### Performance of OPERA in detecting mutations in clinical specimens

To validate the detection performance in real clinical samples, we carried out a prospective clinical study to explore the sensitivity and specificity of OPERA. Forty patients with suspicious NSCLC requiring surgery were recruited, of which there were 39 NSCLC patients and 1 patient with benign disease (Table S7). Ten milliliter blood was collected before surgery, and paired tissue specimens were collected after surgery with pathological confirmation. The majority of NSCLC patients had early-stage disease (stage I, 77%; stage II, 5%), of which 10 patients had a tumor volume of less than 1 cm^3^ (Fig. 4A). DNAs were extracted from tumor tissue, adjacent normal tissue, plasma and blood cell samples correspondingly and tested by OPERA. As a control group, plasma cfDNA samples obtained from 20 healthy individuals were used to determine the mutation cut-off in plasma sample.

**Fig. 4.**
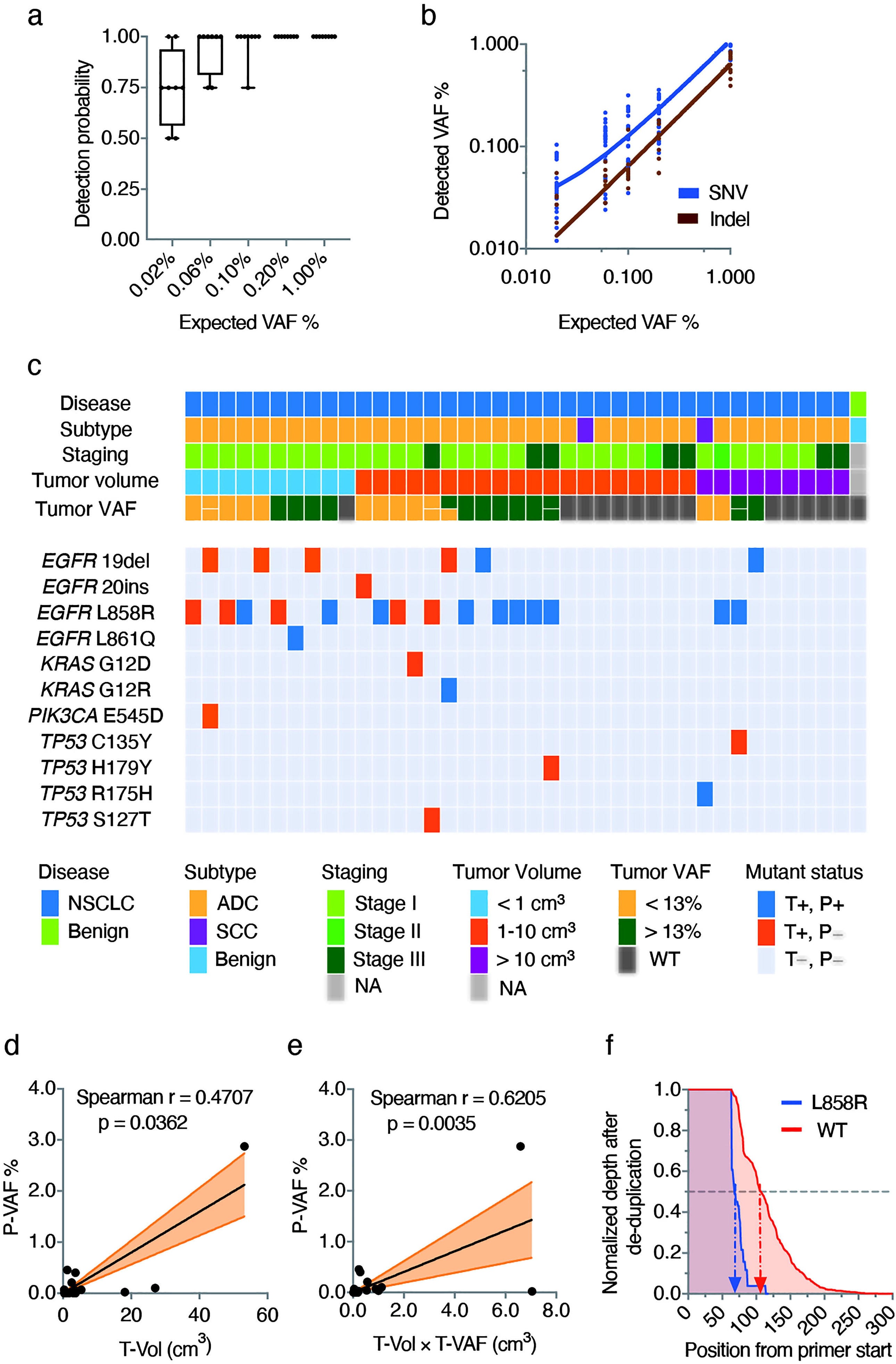
Performance of OPERA in clinical specimens. (a) A box-and-whiskers plot showing the detection probability at different expected variant allelic frequency (VAF) in reference samples. Each experiment was repeated 4 times. (b) Correlation of detected VAF and expected VAF in reference samples. Linear regression of the expected VAF versus the detected VAF of OPERA as classified by single nucleotide variant (SNV) and insertion/deletion (Indel). (c) Detection of somatic mutations in patients. A summary of the somatic mutations detected in tumor tissue and plasma samples and the clinical characteristics of the enrolled patients. Abbreviations: NSCLC, non-small cell lung cancer; ADC, adenocarcinoma; SCC, squamous cell carcinoma; NA, not available; T, tumor tissue; P, plasma. (d-e) Linear regression of (d) the tumor volume versus the plasma VAF and (e) the product of tumor volume and tumor tissue VAF versus the plasma VAF in the single mutation tumors. The correlation is examined by nonparametric Spearman correlation analysis. (f) The length distribution pattern of the *EGFR* L858R mutated and wild-type DNA fragments. The data of all the 10 patients with *EGFR* L858R mutations were summed up in this analysis. The position from primer start is defined as the number of base pairs between the starting position of the DL primer and the end of the paired-end sequencing read.

With a cutoff of 1% VAF and read depth after de-duplication >6500 (Table S8), we found 11 types of somatic mutation in tumor tissue (Fig. 4B). 64.1% (25/39) of NSCLC patients had at least one mutation (included in our 10-hotspot panel) in their tumor tissue (Fig. 4B).The median VAF was 13.16% (ranging from 1.93% to 38.67%) in tumor tissue sample.

The majority of mutation types (9/11) were SNV types. Figure 3 showed that the single-strand error rate of each mutation is significantly different, this would seriously affect the determination of cfDNA mutation. To define the cutoff of each mutation in cfDNA, adjacent normal tissue and blood cell samples, we analyzed substitution type, primer target strand and average single strand error rate (ASSER) (Fig. 3E). We also calculated the max, mean and standard deviation (SD) of VAF in healthy cfDNA samples. Finally, we chose the higher number of the 10× ASSER and mean + 3SD VAF from healthy control group as the cutoff value for each mutation (Table S9). The cutoffs of each SNV mutation were significantly different from 0.001% to 1.27%.

With the specific cutoff for each tumor-derived mutation, half of these mutations (15/30) can be detected in the matched plasma samples (Fig. 4C). The detection sensitivity at sample level was 60.0% (15/25) and the median cfDNA VAF was 0.07% (ranging from 0.01% to 2.88%). The detection specificity was 100% at both mutation level (410/410) and sample level (15/15) (Fig. 4C). Due to a higher read depth after de-duplication (Table S8), OPERA showed a superb performance on *EGFR* SNV detection, with 100% sensitivity on *EGFR* L861Q (1/1) and 66.7% sensitivity on *EGFR* L858R (10/15) (Fig. 4C).

For stage I patients, OPERA detected 50.0% (12/24) of mutations found in matched tumor tissues (57.1% at sample level; 12/21). The plasma VAF of this subgroup ranged from 0.01% to 2.88%. For patients with a tumor volume of around 1 cm^3^, it was reported that the ctDNA VAF was merely 0.008% and practically undetectable [15]. As an ultrasensitive method with mutation-specific cutoffs, OPERA detected 30% (3/10) of mutations in the less than 1 cm^3^ subgroup (33.3% at sample level; 3/9).The plasma VAF of this subgroup ranged from 0.03% to 0.07%. When the tumor volume increased, the plasma detection rate steadily increased. The detection rates of plasma somatic mutations were 53.3% (8/15) and 80.0% (4/5) in the 1-10 cm^3^ subgroup (66.7% at sample level; 8/12; plasma VAF ranged from 0.03% to 1.42%) and the more than 10 cm^3^ subgroup (100% at sample level [4/4]; plasma VAF ranged from 0.01% to 2.88%), respectively.

By plotting a linear regression graph of the tumor volume versus the plasma VAF, a significant positive correlation was observed in the single mutation tumors (Spearman rank analysis, r = 0.470, p = 0.0362; Fig. 4D). Apart from the tumor volume, the tumor tissue VAF is also a key determinant of the plasma VAF. A more significant correlation was observed, when using the product of tumor volume and tumor tissue VAF versus the plasma VAF (Spearman rank analysis r = 0.6205, p = 0.0035; Fig. 4E). These results supported that the plasma somatic mutations detected by OPERA were originated from the matched tumor tissue.

With *EGFR* L858R mutations as an example, the median length of the mutated molecules detected by OPERA was around 70 bp, while that of the wild-type cfDNA was around 105 bp, suggesting that the lengths of ctDNA were significantly shorter (Fig. 4F). This phenomenon is in agreement with previous studies [6,16].

Using the same cutoff for cfDNA samples, the detection specificity of tumor-derived mutations was 100% in both paired adjacent normal tissue and blood cell samples of patients (Fig. S5). Only a few mutations (blood cell sample, 2 of 32; normal tissue, 11 of 32) consistent with the matched tumor tissue were indeed observed with significantly lower VAF (Fig. S5). We speculated that these mutations detected by OPERA in the adjacent normal tissue might originate from occult cancer cells, while those mutations in the blood cell samples might originate from residual ctDNAs or circulating tumor cells.

## Discussion

Tumor tissue positive but ctDNA negative is a relatively common phenomenon [17]. It has been revealed that ctDNA shedding is associated with the baseline tumor size [18] and that a smaller tumor volume results in a lower clonal plasma VAF [15]. Additionally, ctDNAs have been proven to possess a shorter fragment length compared to non-mutant cfDNAs [6,16]. We suggested that the rareness and highly-fragmented nature of ctDNA could be the major obstructions for its detection. Indeed, a recent cross-platform proficiency study applying five commonly used NGS assays concluded that current NGS techniques face challenge in reliably detecting rare ctDNA fragments below the limit of 0.5% VAF [19]. We herein introduce OPERA as a new method for cfDNA detection, especially for highly-fragmented, rare cfDNAs. It has three key features: First, OPERA possesses the highest “theoretical detection ratio”, which represents the proportion for randomly-fragmented molecules to be detected in theory; Secondly, OPERA could detect mutations in short and rare DNA fragments with high library conversion rate; Thirdly, OPERA could suppress single-strand error introduced during sample storage and DNA extraction process. With these advantages, OPERA presented extreme sensitivity for detection of highly-fragmented DNA in plasma sample. In future, this method would also be used in urine or other biological fluids and specimens with seriously damaged DNA. We envision that OPERA could be applied as a powerful molecular diagnostic method in pathogenic microbes detection, early screening/diagnostics/monitoring of cancer, non-invasive prenatal testing, rejection testing for organ transplantation and detection of other disease-associated DNAs.

The strategy of ssDNA library preparation has been shown to be more sensitive to ultra-short cfDNA [20]. A recently published platform, named TARDIS [21], also applied similar strategy to improve the detection sensitivity. However, TARDIS did not solve the by-product problem caused by ligation of primer and adapter during the linear amplification step, therefore, a second nested PCR after ligation was required to distinguish the free primer and linear amplified product, which is not ideal for detecting short fragments due to the longer target length and lower theoretical detection ratio compared to OPERA method.

Previous studies have shown that most of the mutations in plasma come from clonal hematopoiesis [22,23]. However, we did not find more mutations in plasma cfDNA compared with paired saliva samples, in which there supposed not to be affected by clonal hematopoiesis. We speculate that the variation (especially C>T error) we detected with OPERA method mainly comes from single-strand errors that are introduced during sample storage and DNA extraction process, rather than the true variation (including the variation of clonal hematopoiesis) in the sample. These single-strand errors have been reported to be common in FFPE specimens [24]. By designing strand-specific primers, OPERA can dramatically reduce these single-strand errors based on specific substitution types.

Poor concordance of ctDNA mutations with tumor tissue has been a crucial obstacle in early cancer screening [17]. CancerSEEK, as the current most successful mutation detection platform for early cancer screening, tried to promote the sensitivity through multiplex detection with short amplicons of 60-80 bp, yet its sensitivity in stage I lung cancer patients remained disappointing (sensitivity, 43.5%; specificity, 99%) [25]. For OPERA, the sensitivity in stage I lung cancer patients was 57.1% (12/21) with 100% specificity. Compared to a recent study that tried to identify rare ctDNA in stage I-III lung cancers [16], the sensitivity at sample level was similar (59%) even when WES-based customized panel was used, indicating that optimization for multiplexing with more primers might further improve OPERA’s performance.

There are several limitations to this study. First, the simulated samples used in validation experiments cannot perfectly mimic naturally-occurring cfDNAs. For cell line-derived samples, the fragment lengths are not precise due to the restriction of sonication-shearing techniques. While for synthetic DNAs, the genomic coordinates are evenly distributed. Secondly, clinical samples were collected from only 40 patients. Further study with a larger sample size is required to prove the performance of OPERA in early screening of lung cancer.

## Conclusions

OPERA is a novel method for the detection of rare mutations in fragmented DNAs. With the high sensitivity of OPERA in detecting mutations in short DNA fragments with low VAF, it could possibly be applied to early cancer screening.

## Data Availability

All the data associated with this study are present in the paper or the Supplementary Materials.

## List of abbreviations

(ASSER): Average single strand error rate
(cfDNA): Cell-free DNA
(ctDNA): Circulating tumor DNA
(ddPCR): Droplet digital PCR
(DL primer): Dualistic primer
(FFPE): Formalin-fixed paraffin-embedded
(gDNA): Genomic DNA
(Indel): Insertion/deletion
(NGS): Next-generation sequencing
(NSCLC): Non-small cell lung cancer
(OPERA): One-PrimER Amplification
(SNV): Single nucleotide variant
(ssDNA): Single-stranded DNA
(SD): Standard deviation
(UMI): Unique molecular identifier
(VAF): Variant allelic frequency

## Declarations

Ethics approval and consent to participate: The clinical trial was approved by the Ethics Committee of the First Affiliated Hospital of Nanjing Medical University (Reference number: 2019-SR-156).

## Consent for publication

Not applicable.

## Availability of data and materials

All the data associated with this study are present in the paper or the Supplementary Materials.

## Authors’ contributions

LW, GY, ZG, and JL were major contributors in the study conceptualization. LW, QG, RC, ZG, YL, BC, and MR were major contributors in the study methodology. YZ, YY, QG, RH, PZ, QL, and CH were major contributors in the investigation. GY, RC, and GL were major contributors in data analysis and visualization. JL, GY, and WW were responsible for funding acquisition. LW, YY, and ZG were responsible for project administration. GY, WW, and JL were responsible for supervision. RC, GL, and YY were major contributors in writing the manuscript. GY, JL, and WW were major contributors in reviewing and editing the manuscript. All authors read and approved the final manuscript.

## Acknowledgements

We thank all the patients and volunteers that kindly provide their samples to us.

