## Supplementary material for "An ultrasensitive method for detecting mutations from short and rare cell-free DNA": Fig. S1

**TABLE OF CONTENTS**

Fig.S1. Schematic representation of theoretical detection ratio.

Fig. S2. The genomic coordination of the artificial random fragment references and OPERA DL-primer detection system.

Fig. S3. The linear correlation between the variant allelic frequencies as detected by OPERA and that as expected.

Fig. S4. Error rate of OPERA in white blood cell and saliva samples

Fig. S5. Mutation status of paired normal tissue and blood cell samples.

Table S1. The probe regions of CAPP-Seq.

Table S2. Oligonucleotide sequences used for sequencing library preparation.

Table S3. The specific primer sequences of each panel.

Table S4. Ligation efficiency and library conversion rate of unmodified and modified DL-primers.

Table S5. The performance of OPERA in detecting SNV and Indel in cell line samples.

Table S6. Theoretical detection ratio of OPERA.

Table S7. Detailed patient information.

Table S8. Hot-spot depth of de-duplicated reads in 4 types samples.

Table S9. Determination of cutoff value in cfDNA, adjacent normal tissue and blood cell samples.

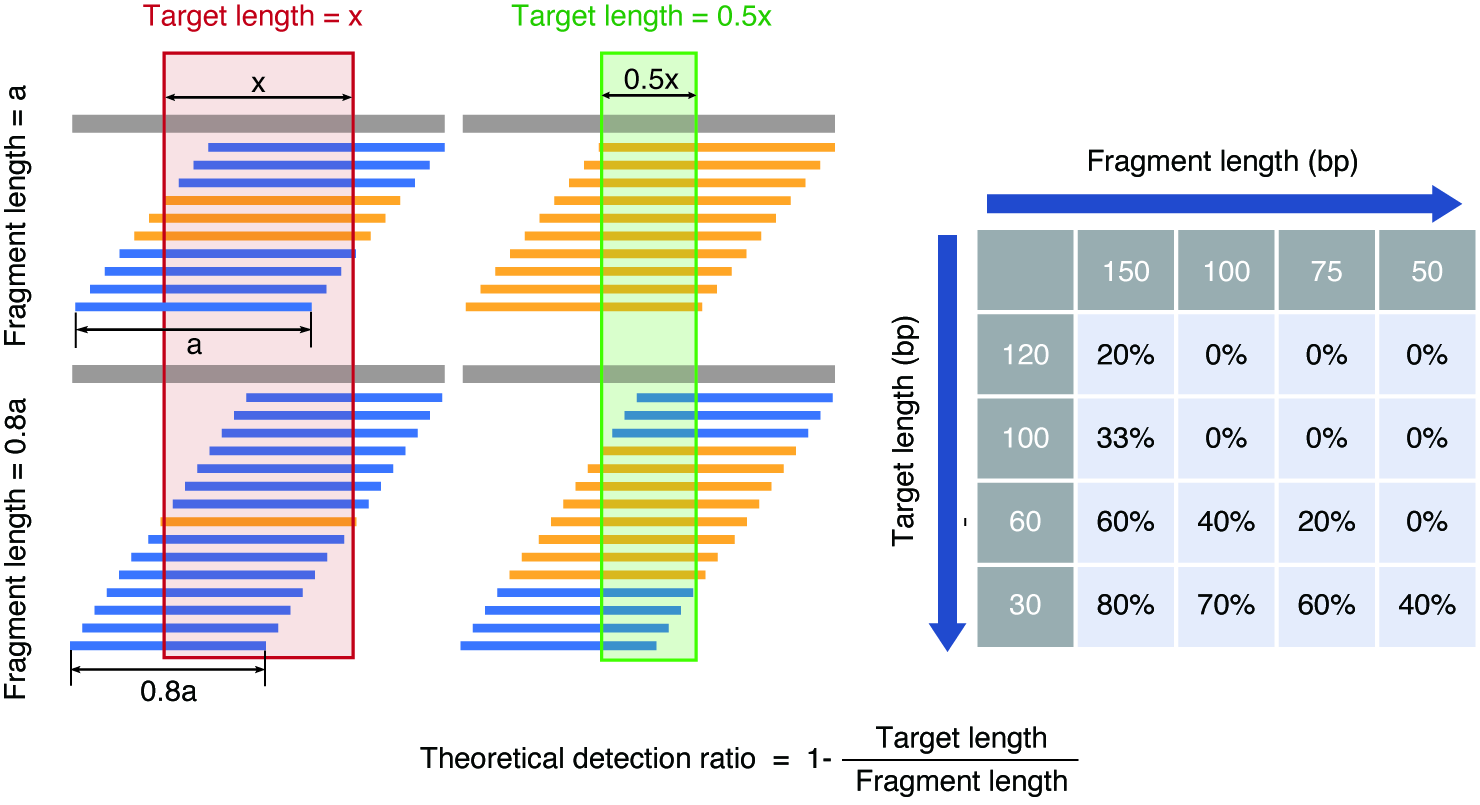

Fig. S1. Schematic representation of theoretical detection ratio. “Target length” represents the sequence length required for detection, “Fragment length” means the length of the DNA fragment. The table on the right shows the theoretical detection ratio at different fragment lengths and different target lengths. The calculation is based on the assumption that fragments with different genomic coordinate are evenly distributed.

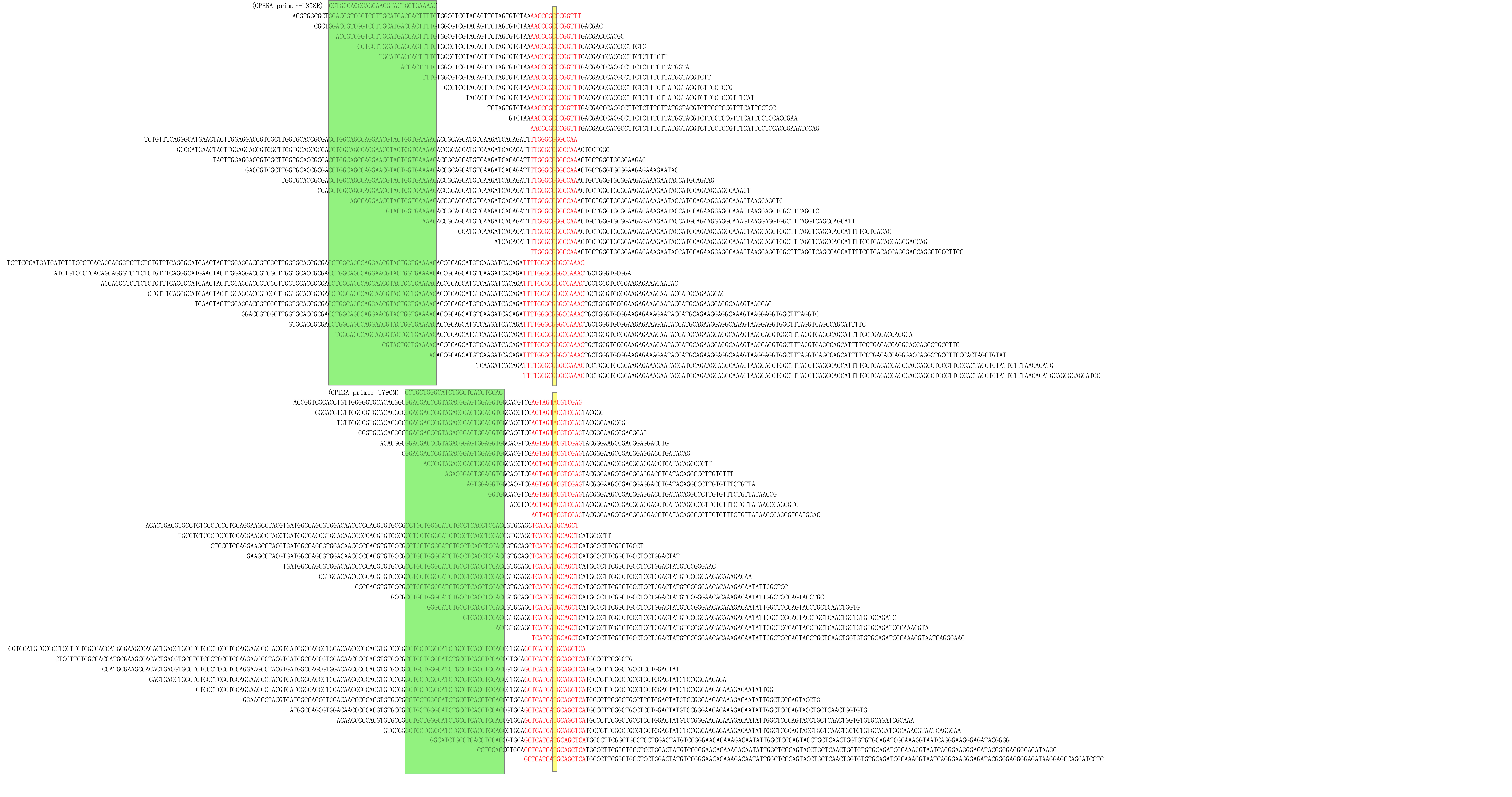

**Fig. S2. The genomic coordination of the artificial random fragment references and OPERA DL-primer detection system.** The sequences of the random fragment references for *EGFR* L858R and T790M mutations with lengths of 80, 120 and 160 bp, with mutation sites indicated in yellow and the OPERA DL-primer sequence indicated in green. The common core regions were indicated in red.

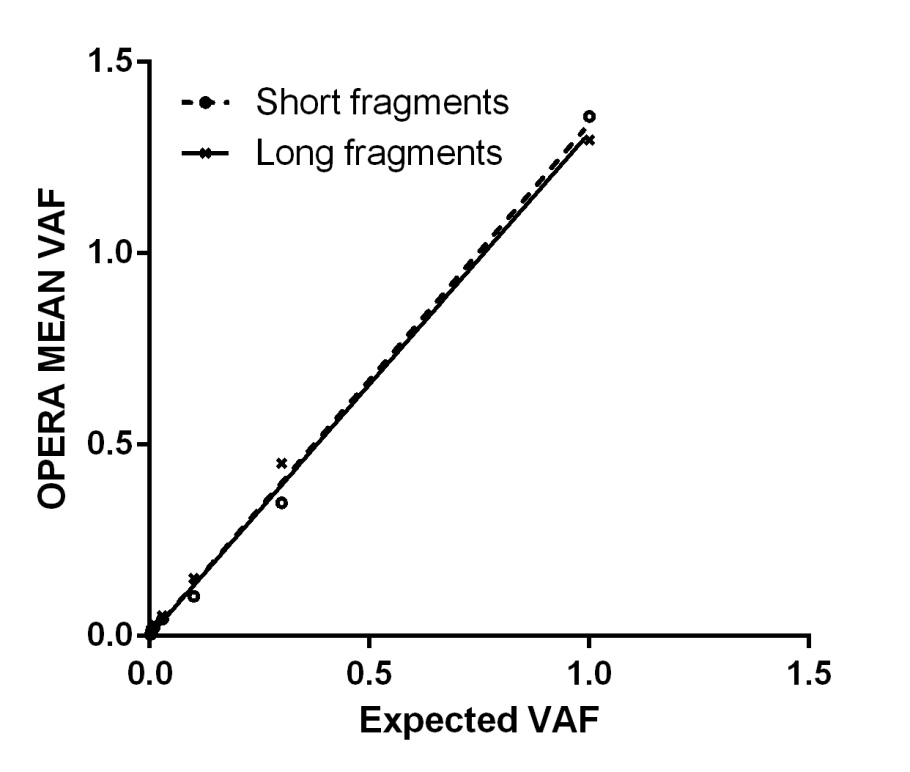

Fig. S3. The linear correlation between the variant allelic frequencies as detected by OPERA and that as expected. The mean VAF as detected by OPERA versus that as expected in cell line-derived DNA fragments with different fragment lengths: dotted line indicated short DNA fragments with 50-150 bp and solid line indicated long DNA fragments with 150-300 bp.

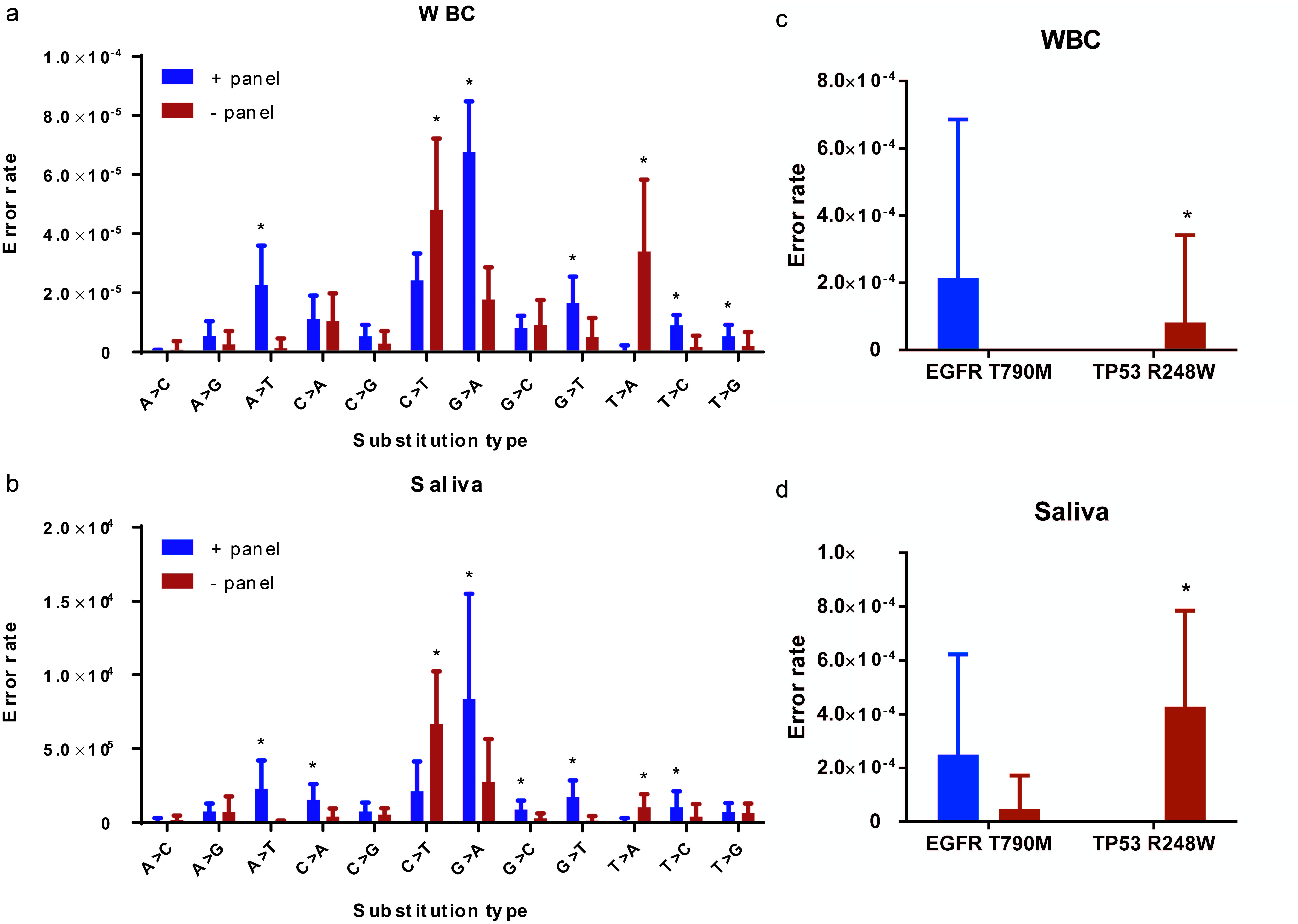

Fig. S4. Error rates of OPERA in white blood cell and saliva samples. Types of error rate of (a and b) all region and (c and d) representative hotspots (*EGFR*-T790M and *TP53*-R248W) as determined by the + panel (bound to plus strand) and the - panel (bound to minus strand) in white blood cell (WBC) and saliva samples, respectively. Errors are distinguished based on the reference sequence of plus strand. The significant difference of error rate detected by the 2 strand-specific panels was examined by paired t test. Asterisk represents *P* value of less than 0.05.

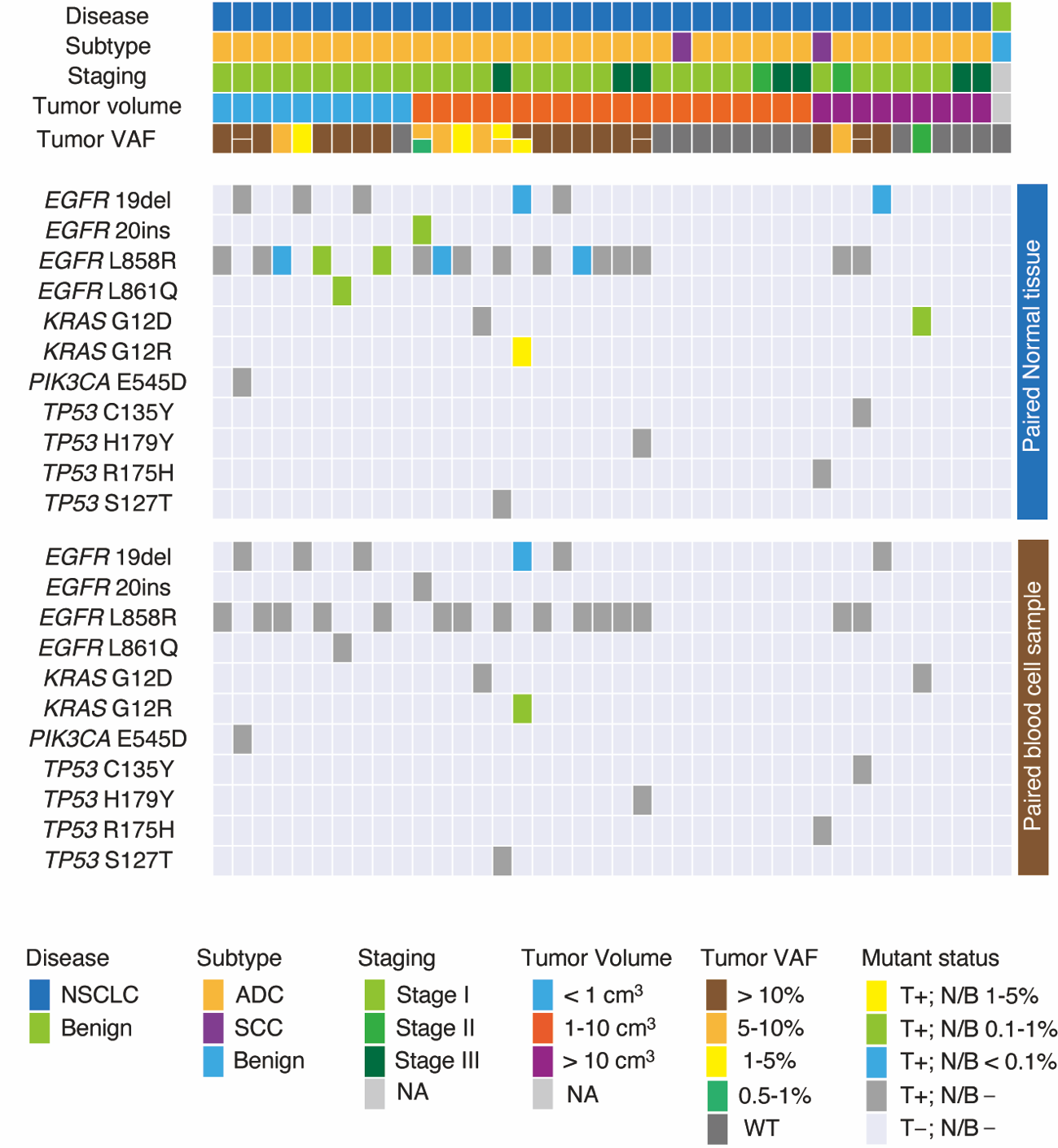

Fig. S5. Mutation status of paired normal tissue and blood cell samples. A summary of the somatic mutations detected in tumor tissue, adjacent normal, and blood cell samples and the clinical characteristics of the enrolled patients. Abbreviations: NSCLC, non-small cell lung cancer; ADC, adenocarcinoma; SCC, squamous cell carcinoma; NA, not available; T, tumor tissue; N, paired normal tissue; B, paired blood cell sample.

**Table S1.** The probe regions of CAPP-Seq.

| chr | from | start |
| --- | --- | --- |
| chr3 | 179218209 | 179218334 |
| chr7 | 140753274 | 140753393 |
| chr1 | 114713799 | 114713978 |
| chr7 | 55191718 | 55191874 |
| chr7 | 55181292 | 55181478 |
| chr7 | 55174721 | 55174820 |
| chr17 | 39724725 | 39724911 |
| chr12 | 25245273 | 25245384 |
| chr12 | 25227233 | 25227412 |
| chr17 | 7674858 | 7674971 |
| chr1 | 11107484 | 11107500 |
| chr1 | 11108180 | 11108286 |
| chr1 | 11109289 | 11109370 |
| chr1 | 11109648 | 11109729 |
| chr1 | 11112851 | 11112917 |
| chr1 | 11114317 | 11114453 |
| chr1 | 11114812 | 11114887 |
| chr1 | 11115395 | 11115468 |
| chr1 | 11117003 | 11117086 |
| chr1 | 11121245 | 11121368 |
| chr1 | 11121978 | 11122126 |
| chr1 | 11124497 | 11124633 |
| chr1 | 11126621 | 11126796 |
| chr1 | 11127009 | 11127144 |
| chr1 | 11127623 | 11127806 |
| chr1 | 11128003 | 11128126 |
| chr1 | 11128453 | 11128552 |
| chr1 | 11128854 | 11128951 |
| chr1 | 11129737 | 11129838 |
| chr1 | 11130528 | 11130777 |
| chr1 | 11133079 | 11133197 |
| chr1 | 11134350 | 11134466 |
| chr1 | 11139303 | 11139435 |
| chr1 | 11139532 | 11139658 |
| chr1 | 11144647 | 11144755 |
| chr1 | 11144967 | 11145045 |
| chr1 | 11146675 | 11146791 |
| chr1 | 11150125 | 11150226 |
| chr1 | 11157151 | 11157291 |
| chr1 | 11167441 | 11167517 |
| chr1 | 11199257 | 11199403 |
| chr1 | 11199540 | 11199703 |
| chr1 | 11204560 | 11204703 |
| chr1 | 11209311 | 11209458 |
| chr1 | 11210813 | 11210906 |
| chr1 | 11212311 | 11212474 |
| chr1 | 11212795 | 11212908 |
| chr1 | 11213398 | 11213566 |
| chr1 | 11216147 | 11216234 |
| chr1 | 11228667 | 11228918 |
| chr1 | 11230924 | 11231054 |
| chr1 | 11231299 | 11231434 |
| chr1 | 11232435 | 11232528 |
| chr1 | 11233397 | 11233487 |
| chr1 | 11234142 | 11234265 |
| chr1 | 11237842 | 11238048 |
| chr1 | 11238401 | 11238617 |
| chr1 | 11240302 | 11240547 |
| chr1 | 11241552 | 11241681 |
| chr1 | 11243113 | 11243300 |
| chr1 | 11247624 | 11247733 |
| chr1 | 11247818 | 11248094 |
| chr1 | 11253838 | 11253973 |
| chr1 | 11255991 | 11256192 |
| chr1 | 11256932 | 11257165 |
| chr1 | 11258484 | 11258593 |
| chr1 | 11259247 | 11259409 |
| chr1 | 45329305 | 45329437 |
| chr1 | 45330515 | 45330557 |
| chr1 | 45331181 | 45331334 |
| chr1 | 45331419 | 45331556 |
| chr1 | 45331660 | 45331849 |
| chr1 | 45332022 | 45332086 |
| chr1 | 45332165 | 45332310 |
| chr1 | 45332390 | 45332488 |
| chr1 | 45332573 | 45332687 |
| chr1 | 45332762 | 45332834 |
| chr1 | 45332917 | 45332959 |
| chr1 | 45333096 | 45333170 |
| chr1 | 45333284 | 45333324 |
| chr1 | 45333412 | 45333594 |
| chr1 | 45334390 | 45334511 |
| chr1 | 45340218 | 45340254 |
| chr1 | 46248405 | 46248408 |
| chr1 | 46248511 | 46248598 |
| chr1 | 46249999 | 46250119 |
| chr1 | 46258685 | 46258746 |
| chr1 | 46259963 | 46260099 |
| chr1 | 46260541 | 46260611 |
| chr1 | 46260726 | 46261015 |
| chr1 | 46261260 | 46261385 |
| chr1 | 46267458 | 46267609 |
| chr1 | 46270658 | 46270785 |
| chr1 | 46272465 | 46272540 |
| chr1 | 46272671 | 46272802 |
| chr1 | 46273354 | 46273465 |
| chr1 | 46273623 | 46273747 |
| chr1 | 46274137 | 46274216 |
| chr1 | 46274537 | 46274717 |
| chr1 | 46277816 | 46277980 |
| chr1 | 46278071 | 46278282 |
| chr1 | 114708534 | 114708654 |
| chr1 | 114709568 | 114709728 |
| chr1 | 114716049 | 114716160 |
| chr1 | 156815829 | 156815838 |
| chr1 | 156842080 | 156842193 |
| chr1 | 156860934 | 156861146 |
| chr1 | 156864353 | 156864428 |
| chr1 | 156864727 | 156866978 |
| chr1 | 156868103 | 156868249 |
| chr1 | 156868504 | 156868647 |
| chr1 | 156871622 | 156872702 |
| chr1 | 156872942 | 156877082 |
| chr1 | 156877142 | 156879362 |
| chr1 | 156879998 | 156880157 |
| chr1 | 156881456 | 156881642 |
| chr1 | 162719063 | 162719145 |
| chr1 | 162753094 | 162753197 |
| chr1 | 162754623 | 162754855 |
| chr1 | 162755155 | 162755303 |
| chr1 | 162755663 | 162755769 |
| chr1 | 162759795 | 162759979 |
| chr1 | 162761210 | 162761454 |
| chr1 | 162766000 | 162766063 |
| chr1 | 162767228 | 162767359 |
| chr1 | 162770301 | 162770512 |
| chr1 | 162772023 | 162772247 |
| chr1 | 162773468 | 162773596 |
| chr1 | 162775651 | 162775843 |
| chr1 | 162776135 | 162776370 |
| chr1 | 162778579 | 162778729 |
| chr1 | 162780111 | 162780246 |
| chr10 | 43077258 | 43077331 |
| chr10 | 43100458 | 43100722 |
| chr10 | 43102341 | 43102629 |
| chr10 | 43104951 | 43105193 |
| chr10 | 43106375 | 43106571 |
| chr10 | 43109030 | 43116731 |
| chr10 | 43118372 | 43118480 |
| chr10 | 43119530 | 43119745 |
| chr10 | 43120080 | 43120203 |
| chr10 | 43121945 | 43122016 |
| chr10 | 43123670 | 43123808 |
| chr10 | 43124882 | 43124982 |
| chr10 | 43126574 | 43126754 |
| chr10 | 43128111 | 43128269 |
| chr10 | 87864469 | 87864548 |
| chr10 | 87894024 | 87894109 |
| chr10 | 87925442 | 87925627 |
| chr10 | 87931045 | 87931089 |
| chr10 | 87933012 | 87933251 |
| chr10 | 87952117 | 87952259 |
| chr10 | 87957852 | 87958019 |
| chr10 | 87960893 | 87961118 |
| chr10 | 87965286 | 87965472 |
| chr10 | 121479580 | 121479670 |
| chr10 | 121479856 | 121480021 |
| chr10 | 121481100 | 121481161 |
| chr10 | 121482171 | 121482177 |
| chr10 | 121483697 | 121483803 |
| chr10 | 121485394 | 121485532 |
| chr10 | 121487353 | 121487424 |
| chr10 | 121487990 | 121488113 |
| chr10 | 121496531 | 121496722 |
| chr10 | 121498494 | 121498605 |
| chr10 | 121500825 | 121500947 |
| chr10 | 121503789 | 121503941 |
| chr10 | 121504071 | 121504132 |
| chr10 | 121515116 | 121515319 |
| chr10 | 121516546 | 121516607 |
| chr10 | 121517318 | 121517463 |
| chr10 | 121518681 | 121518829 |
| chr10 | 121519978 | 121520169 |
| chr10 | 121526154 | 121526194 |
| chr10 | 121526728 | 121526752 |
| chr10 | 121538591 | 121538715 |
| chr10 | 121551289 | 121551459 |
| chr10 | 121552153 | 121552214 |
| chr10 | 121558186 | 121558247 |
| chr10 | 121564501 | 121564579 |
| chr10 | 121565437 | 121565704 |
| chr10 | 121566210 | 121566271 |
| chr10 | 121569884 | 121569945 |
| chr10 | 121579109 | 121579170 |
| chr10 | 121586571 | 121586632 |
| chr10 | 121593708 | 121593817 |
| chr11 | 532635 | 532755 |
| chr11 | 533295 | 533358 |
| chr11 | 533452 | 533612 |
| chr11 | 533765 | 533944 |
| chr11 | 534211 | 534322 |
| chr11 | 69641313 | 69641511 |
| chr11 | 69643030 | 69643246 |
| chr11 | 69643831 | 69643991 |
| chr11 | 69647993 | 69648142 |
| chr11 | 69651117 | 69651282 |
| chr11 | 108227624 | 108227696 |
| chr11 | 108227775 | 108227888 |
| chr11 | 108229177 | 108229331 |
| chr11 | 108235669 | 108235834 |
| chr11 | 108243952 | 108244118 |
| chr11 | 108244787 | 108245026 |
| chr11 | 108246963 | 108247127 |
| chr11 | 108248932 | 108249102 |
| chr11 | 108250700 | 108251072 |
| chr11 | 108251836 | 108252031 |
| chr11 | 108252816 | 108252912 |
| chr11 | 108253813 | 108254039 |
| chr11 | 108256214 | 108256340 |
| chr11 | 108257480 | 108257606 |
| chr11 | 108258985 | 108259075 |
| chr11 | 108267170 | 108267342 |
| chr11 | 108268409 | 108268609 |
| chr11 | 108271063 | 108271146 |
| chr11 | 108271250 | 108271406 |
| chr11 | 108272531 | 108272607 |
| chr11 | 108272721 | 108272852 |
| chr11 | 108279490 | 108279608 |
| chr11 | 108280994 | 108281168 |
| chr11 | 108282709 | 108282879 |
| chr11 | 108284226 | 108284473 |
| chr11 | 108287599 | 108287715 |
| chr11 | 108288976 | 108289103 |
| chr11 | 108289601 | 108289801 |
| chr11 | 108292618 | 108292793 |
| chr11 | 108293312 | 108293477 |
| chr11 | 108294926 | 108295059 |
| chr11 | 108297286 | 108297382 |
| chr11 | 108299713 | 108299885 |
| chr11 | 108301647 | 108301789 |
| chr11 | 108302852 | 108303029 |
| chr11 | 108304674 | 108304852 |
| chr11 | 108307896 | 108307984 |
| chr11 | 108310159 | 108310315 |
| chr11 | 108312410 | 108312498 |
| chr11 | 108315822 | 108315911 |
| chr11 | 108316010 | 108316113 |
| chr11 | 108317372 | 108317521 |
| chr11 | 108319953 | 108320058 |
| chr11 | 108321300 | 108321420 |
| chr11 | 108325309 | 108325544 |
| chr11 | 108326057 | 108326225 |
| chr11 | 108327644 | 108327758 |
| chr11 | 108329020 | 108329238 |
| chr11 | 108330213 | 108330421 |
| chr11 | 108331443 | 108331557 |
| chr11 | 108331878 | 108332037 |
| chr11 | 108332761 | 108332900 |
| chr11 | 108333885 | 108333968 |
| chr11 | 108334968 | 108335109 |
| chr11 | 108335844 | 108335961 |
| chr11 | 108343221 | 108343371 |
| chr11 | 108345742 | 108345908 |
| chr11 | 108347278 | 108347365 |
| chr11 | 108353765 | 108353880 |
| chr11 | 108354810 | 108354874 |
| chr11 | 108365081 | 108365218 |
| chr11 | 108365324 | 108365508 |
| chr11 | 125626768 | 125626833 |
| chr11 | 125627606 | 125627830 |
| chr11 | 125629231 | 125629296 |
| chr11 | 125629390 | 125629460 |
| chr11 | 125633162 | 125633351 |
| chr11 | 125635428 | 125635533 |
| chr11 | 125637448 | 125637544 |
| chr11 | 125643791 | 125643900 |
| chr11 | 125644090 | 125644268 |
| chr11 | 125644511 | 125644643 |
| chr11 | 125653745 | 125653847 |
| chr11 | 125655224 | 125655320 |
| chr12 | 25209794 | 25209911 |
| chr12 | 25215440 | 25215560 |
| chr12 | 25225613 | 25225773 |
| chr12 | 56084973 | 56085201 |
| chr12 | 56086509 | 56086676 |
| chr12 | 56087773 | 56087933 |
| chr12 | 56087999 | 56088182 |
| chr12 | 56088521 | 56088676 |
| chr12 | 56088726 | 56088888 |
| chr12 | 56093323 | 56093570 |
| chr12 | 56094380 | 56094576 |
| chr12 | 56095643 | 56095826 |
| chr12 | 56097023 | 56097250 |
| chr12 | 56097763 | 56097960 |
| chr12 | 56098737 | 56098925 |
| chr12 | 56101039 | 56101381 |
| chr12 | 56101507 | 56102075 |
| chr12 | 57748524 | 57748617 |
| chr12 | 57749181 | 57749317 |
| chr12 | 57749453 | 57749504 |
| chr12 | 57750655 | 57750765 |
| chr12 | 57750922 | 57751090 |
| chr12 | 57751206 | 57751342 |
| chr12 | 57751499 | 57751717 |
| chr12 | 132624696 | 132624810 |
| chr12 | 132624904 | 132624994 |
| chr12 | 132625644 | 132625770 |
| chr12 | 132626116 | 132626317 |
| chr12 | 132632314 | 132632508 |
| chr12 | 132632663 | 132632795 |
| chr12 | 132634185 | 132634378 |
| chr12 | 132635891 | 132636024 |
| chr12 | 132638013 | 132638139 |
| chr12 | 132639124 | 132639298 |
| chr12 | 132641646 | 132641851 |
| chr12 | 132642176 | 132642397 |
| chr12 | 132642505 | 132642729 |
| chr12 | 132642819 | 132642996 |
| chr12 | 132643223 | 132643330 |
| chr12 | 132643406 | 132643560 |
| chr12 | 132643836 | 132643977 |
| chr12 | 132648928 | 132649072 |
| chr12 | 132649305 | 132649515 |
| chr12 | 132649676 | 132649889 |
| chr12 | 132657135 | 132657258 |
| chr12 | 132657348 | 132657429 |
| chr12 | 132657867 | 132657970 |
| chr12 | 132659294 | 132659509 |
| chr12 | 132660968 | 132661164 |
| chr12 | 132661526 | 132661684 |
| chr12 | 132664003 | 132664148 |
| chr12 | 132664369 | 132664462 |
| chr12 | 132665301 | 132665450 |
| chr12 | 132667502 | 132667648 |
| chr12 | 132668355 | 132668502 |
| chr12 | 132668634 | 132668737 |
| chr12 | 132668810 | 132668939 |
| chr12 | 132672214 | 132672322 |
| chr12 | 132672626 | 132672839 |
| chr12 | 132673163 | 132673277 |
| chr12 | 132673574 | 132673707 |
| chr12 | 132675397 | 132675517 |
| chr12 | 132675734 | 132675820 |
| chr12 | 132676093 | 132676204 |
| chr12 | 132676545 | 132676653 |
| chr12 | 132677362 | 132677443 |
| chr12 | 132677577 | 132677719 |
| chr12 | 132679496 | 132679651 |
| chr12 | 132679953 | 132680046 |
| chr12 | 132680177 | 132680222 |
| chr12 | 132680606 | 132680687 |
| chr12 | 132681137 | 132681279 |
| chr12 | 132687253 | 132687315 |
| chr13 | 32316460 | 32316527 |
| chr13 | 32319076 | 32319325 |
| chr13 | 32325075 | 32325184 |
| chr13 | 32326100 | 32326150 |
| chr13 | 32326241 | 32326282 |
| chr13 | 32326498 | 32326613 |
| chr13 | 32329362 | 32329572 |
| chr13 | 32330918 | 32331030 |
| chr13 | 32332271 | 32333387 |
| chr13 | 32336264 | 32341196 |
| chr13 | 32344557 | 32344653 |
| chr13 | 32346826 | 32346896 |
| chr13 | 32354860 | 32355288 |
| chr13 | 32356427 | 32356609 |
| chr13 | 32357741 | 32357929 |
| chr13 | 32362522 | 32362693 |
| chr13 | 32363178 | 32363533 |
| chr13 | 32370401 | 32370557 |
| chr13 | 32370955 | 32371100 |
| chr13 | 32376669 | 32376791 |
| chr13 | 32379316 | 32379515 |
| chr13 | 32379749 | 32379913 |
| chr13 | 32380006 | 32380145 |
| chr13 | 32394688 | 32394933 |
| chr13 | 32396897 | 32397044 |
| chr13 | 32398161 | 32398770 |
| chr14 | 67823543 | 67823627 |
| chr14 | 67825463 | 67825577 |
| chr14 | 67835079 | 67835196 |
| chr14 | 67865002 | 67865139 |
| chr14 | 67885868 | 67885988 |
| chr14 | 67887020 | 67887204 |
| chr14 | 68291883 | 68291980 |
| chr14 | 68411423 | 68411527 |
| chr14 | 68468171 | 68468250 |
| chr14 | 68477647 | 68477664 |
| chr14 | 68611005 | 68611247 |
| chr14 | 104770340 | 104770420 |
| chr14 | 104770744 | 104770847 |
| chr14 | 104772364 | 104772452 |
| chr14 | 104772877 | 104773092 |
| chr14 | 104773250 | 104773379 |
| chr14 | 104773454 | 104773580 |
| chr14 | 104773911 | 104773980 |
| chr14 | 104774937 | 104775003 |
| chr14 | 104775075 | 104775207 |
| chr14 | 104775651 | 104775799 |
| chr14 | 104776658 | 104776770 |
| chr14 | 104780087 | 104780216 |
| chr14 | 104792597 | 104792643 |
| chr15 | 66387347 | 66387427 |
| chr15 | 66435026 | 66435237 |
| chr15 | 66436745 | 66436892 |
| chr15 | 66443279 | 66443357 |
| chr15 | 66444655 | 66444707 |
| chr15 | 66481754 | 66481879 |
| chr15 | 66484989 | 66485191 |
| chr15 | 66487227 | 66487292 |
| chr15 | 66489214 | 66489276 |
| chr15 | 66489717 | 66489763 |
| chr15 | 66490501 | 66490615 |
| chr15 | 87876934 | 87877120 |
| chr15 | 87880269 | 87880428 |
| chr15 | 87885613 | 87885815 |
| chr15 | 87916545 | 87916560 |
| chr15 | 87929190 | 87929434 |
| chr15 | 87933011 | 87933184 |
| chr15 | 87940622 | 87940753 |
| chr15 | 87979344 | 87979463 |
| chr15 | 87981225 | 87981360 |
| chr15 | 88032856 | 88033045 |
| chr15 | 88126270 | 88126373 |
| chr15 | 88127161 | 88127226 |
| chr15 | 88128710 | 88128734 |
| chr15 | 88135100 | 88135397 |
| chr15 | 88135898 | 88136040 |
| chr15 | 88136466 | 88136609 |
| chr15 | 88137403 | 88137561 |
| chr15 | 88147334 | 88147403 |
| chr15 | 88183417 | 88183489 |
| chr15 | 88184224 | 88184299 |
| chr15 | 88255905 | 88256153 |
| chr15 | 90085253 | 90085407 |
| chr15 | 90088358 | 90088767 |
| chr16 | 2039923 | 2040047 |
| chr16 | 2040132 | 2040238 |
| chr16 | 2043566 | 2043726 |
| chr16 | 2044629 | 2044800 |
| chr16 | 2046127 | 2046366 |
| chr16 | 2047708 | 2047823 |
| chr16 | 23603458 | 23603669 |
| chr16 | 23607863 | 23608012 |
| chr16 | 23614003 | 23614091 |
| chr16 | 23621361 | 23621478 |
| chr16 | 23622968 | 23623130 |
| chr16 | 23624008 | 23624094 |
| chr16 | 23626235 | 23626397 |
| chr16 | 23629203 | 23629275 |
| chr16 | 23629639 | 23630469 |
| chr16 | 23634861 | 23636334 |
| chr16 | 23637849 | 23637952 |
| chr16 | 23638069 | 23638129 |
| chr16 | 23641109 | 23641157 |
| chr17 | 7669608 | 7669690 |
| chr17 | 7670608 | 7670715 |
| chr17 | 7673218 | 7673266 |
| chr17 | 7673306 | 7673339 |
| chr17 | 7673534 | 7673608 |
| chr17 | 7673700 | 7673837 |
| chr17 | 7674180 | 7674290 |
| chr17 | 7675052 | 7675236 |
| chr17 | 7675993 | 7676272 |
| chr17 | 7676381 | 7676403 |
| chr17 | 7676520 | 7676594 |
| chr17 | 31095309 | 31095369 |
| chr17 | 31155982 | 31156126 |
| chr17 | 31159009 | 31159093 |
| chr17 | 31163185 | 31163376 |
| chr17 | 31169890 | 31169997 |
| chr17 | 31181421 | 31181489 |
| chr17 | 31181709 | 31181785 |
| chr17 | 31182507 | 31182665 |
| chr17 | 31200421 | 31200595 |
| chr17 | 31201036 | 31201159 |
| chr17 | 31201410 | 31201485 |
| chr17 | 31206239 | 31206371 |
| chr17 | 31214450 | 31214585 |
| chr17 | 31219004 | 31219118 |
| chr17 | 31221849 | 31221990 |
| chr17 | 31223443 | 31223567 |
| chr17 | 31225094 | 31225250 |
| chr17 | 31226434 | 31226684 |
| chr17 | 31227217 | 31227291 |
| chr17 | 31227522 | 31227606 |
| chr17 | 31229024 | 31229465 |
| chr17 | 31229834 | 31229974 |
| chr17 | 31230259 | 31230382 |
| chr17 | 31230841 | 31230925 |
| chr17 | 31232072 | 31232189 |
| chr17 | 31232699 | 31232881 |
| chr17 | 31233001 | 31233213 |
| chr17 | 31235610 | 31235772 |
| chr17 | 31235917 | 31236021 |
| chr17 | 31248983 | 31249119 |
| chr17 | 31252937 | 31253000 |
| chr17 | 31258343 | 31258502 |
| chr17 | 31259031 | 31259129 |
| chr17 | 31260368 | 31260515 |
| chr17 | 31261710 | 31261857 |
| chr17 | 31265228 | 31265339 |
| chr17 | 31325819 | 31326252 |
| chr17 | 31327498 | 31327839 |
| chr17 | 31330295 | 31330498 |
| chr17 | 31334837 | 31335031 |
| chr17 | 31336332 | 31336473 |
| chr17 | 31336634 | 31336914 |
| chr17 | 31337367 | 31337582 |
| chr17 | 31337818 | 31337880 |
| chr17 | 31338024 | 31338139 |
| chr17 | 31338703 | 31338805 |
| chr17 | 31340504 | 31340645 |
| chr17 | 31343008 | 31343135 |
| chr17 | 31349119 | 31349251 |
| chr17 | 31350182 | 31350318 |
| chr17 | 31352256 | 31352414 |
| chr17 | 31356459 | 31356582 |
| chr17 | 31356959 | 31357090 |
| chr17 | 31357268 | 31357369 |
| chr17 | 31358479 | 31358622 |
| chr17 | 31358968 | 31359015 |
| chr17 | 31360486 | 31360703 |
| chr17 | 31374012 | 31374155 |
| chr17 | 35100952 | 35101036 |
| chr17 | 35101200 | 35101365 |
| chr17 | 35103253 | 35103324 |
| chr17 | 35103453 | 35103544 |
| chr17 | 35106385 | 35106481 |
| chr17 | 35106987 | 35107122 |
| chr17 | 35107365 | 35107447 |
| chr17 | 35116858 | 35117037 |
| chr17 | 35118500 | 35118619 |
| chr17 | 35119110 | 35119172 |
| chr17 | 35119531 | 35119613 |
| chr17 | 39462071 | 39463117 |
| chr17 | 39470878 | 39471763 |
| chr17 | 39490556 | 39490733 |
| chr17 | 39492750 | 39492890 |
| chr17 | 39494523 | 39494694 |
| chr17 | 39501249 | 39501439 |
| chr17 | 39509704 | 39509761 |
| chr17 | 39511528 | 39511630 |
| chr17 | 39515730 | 39515808 |
| chr17 | 39517439 | 39517556 |
| chr17 | 39519955 | 39520087 |
| chr17 | 39524673 | 39524885 |
| chr17 | 39525863 | 39526316 |
| chr17 | 39530603 | 39531316 |
| chr17 | 39699559 | 39699587 |
| chr17 | 39700238 | 39700311 |
| chr17 | 39702394 | 39702455 |
| chr17 | 39705434 | 39705495 |
| chr17 | 39706989 | 39707141 |
| chr17 | 39708320 | 39708534 |
| chr17 | 39709317 | 39709452 |
| chr17 | 39709812 | 39709881 |
| chr17 | 39710085 | 39710201 |
| chr17 | 39710339 | 39710481 |
| chr17 | 39711927 | 39712047 |
| chr17 | 39712321 | 39712448 |
| chr17 | 39714094 | 39714155 |
| chr17 | 39715285 | 39715359 |
| chr17 | 39715445 | 39715536 |
| chr17 | 39715739 | 39715939 |
| chr17 | 39716300 | 39716433 |
| chr17 | 39716514 | 39716605 |
| chr17 | 39717319 | 39717484 |
| chr17 | 39719786 | 39719834 |
| chr17 | 39720551 | 39720612 |
| chr17 | 39723318 | 39723457 |
| chr17 | 39723537 | 39723660 |
| chr17 | 39723911 | 39724010 |
| chr17 | 39725048 | 39725204 |
| chr17 | 39725326 | 39725402 |
| chr17 | 39725706 | 39725853 |
| chr17 | 39726561 | 39726659 |
| chr17 | 39726814 | 39727003 |
| chr17 | 39727294 | 39727547 |
| chr17 | 39727688 | 39728044 |
| chr17 | 43045677 | 43045802 |
| chr17 | 43047642 | 43047703 |
| chr17 | 43049120 | 43049194 |
| chr17 | 43051062 | 43051117 |
| chr17 | 43057051 | 43057135 |
| chr17 | 43063332 | 43063373 |
| chr17 | 43063873 | 43063951 |
| chr17 | 43067607 | 43067695 |
| chr17 | 43070927 | 43071238 |
| chr17 | 43074330 | 43074521 |
| chr17 | 43076487 | 43076614 |
| chr17 | 43079333 | 43079399 |
| chr17 | 43082403 | 43082575 |
| chr17 | 43090943 | 43091032 |
| chr17 | 43091434 | 43094860 |
| chr17 | 43095845 | 43095922 |
| chr17 | 43097243 | 43097289 |
| chr17 | 43099774 | 43099880 |
| chr17 | 43104121 | 43104261 |
| chr17 | 43104867 | 43104956 |
| chr17 | 43106455 | 43106533 |
| chr17 | 43115725 | 43115779 |
| chr17 | 43124016 | 43124096 |
| chr17 | 58692643 | 58692788 |
| chr17 | 58694930 | 58695193 |
| chr17 | 58696692 | 58696859 |
| chr17 | 58703195 | 58703329 |
| chr17 | 58709858 | 58709990 |
| chr17 | 58720745 | 58720812 |
| chr17 | 58724039 | 58724100 |
| chr17 | 58732483 | 58732544 |
| chr17 | 58734117 | 58734222 |
| chr17 | 61683295 | 61684140 |
| chr17 | 61685835 | 61686165 |
| chr17 | 61693429 | 61693512 |
| chr17 | 61715950 | 61716063 |
| chr17 | 61743012 | 61743134 |
| chr17 | 61744431 | 61744591 |
| chr17 | 61776400 | 61776562 |
| chr17 | 61780260 | 61780401 |
| chr17 | 61780839 | 61781005 |
| chr17 | 61784269 | 61784424 |
| chr17 | 61793596 | 61793729 |
| chr17 | 61799099 | 61799299 |
| chr17 | 61801252 | 61801474 |
| chr17 | 61808466 | 61808757 |
| chr17 | 61847100 | 61847220 |
| chr17 | 61849128 | 61849256 |
| chr17 | 61857057 | 61857231 |
| chr17 | 61859795 | 61859907 |
| chr17 | 61861446 | 61861539 |
| chr18 | 51047046 | 51047295 |
| chr18 | 51048685 | 51048860 |
| chr18 | 51049294 | 51049324 |
| chr18 | 51054780 | 51054993 |
| chr18 | 51058124 | 51058244 |
| chr18 | 51058339 | 51058456 |
| chr18 | 51059865 | 51059916 |
| chr18 | 51065422 | 51065606 |
| chr18 | 51067018 | 51067187 |
| chr18 | 51076637 | 51076776 |
| chr18 | 51078255 | 51078467 |
| chr19 | 1206913 | 1207203 |
| chr19 | 1218416 | 1218500 |
| chr19 | 1219323 | 1219413 |
| chr19 | 1220372 | 1220505 |
| chr19 | 1220580 | 1220717 |
| chr19 | 1221212 | 1221340 |
| chr19 | 1221948 | 1222006 |
| chr19 | 1222984 | 1223172 |
| chr19 | 1226453 | 1226647 |
| chr19 | 4100997 | 4101163 |
| chr19 | 4102354 | 4102473 |
| chr19 | 4110487 | 4110675 |
| chr19 | 4117397 | 4117649 |
| chr19 | 10486651 | 10486818 |
| chr19 | 10489191 | 10489368 |
| chr19 | 10489647 | 10489853 |
| chr19 | 10491576 | 10492262 |
| chr19 | 10499394 | 10500033 |
| chr19 | 50398851 | 50399053 |
| chr19 | 50399370 | 50399484 |
| chr19 | 50401777 | 50401924 |
| chr19 | 50401998 | 50402124 |
| chr19 | 50402204 | 50402373 |
| chr19 | 50402453 | 50402535 |
| chr19 | 50402611 | 50402741 |
| chr19 | 50403052 | 50403219 |
| chr19 | 50403492 | 50403597 |
| chr19 | 50406181 | 50406322 |
| chr19 | 50406406 | 50406517 |
| chr19 | 50406982 | 50407174 |
| chr19 | 50407326 | 50407415 |
| chr19 | 50408706 | 50408901 |
| chr19 | 50409121 | 50409235 |
| chr19 | 50409518 | 50409666 |
| chr19 | 50413425 | 50413521 |
| chr19 | 50413741 | 50413879 |
| chr19 | 50414814 | 50414990 |
| chr19 | 50415437 | 50415590 |
| chr19 | 50415723 | 50415826 |
| chr19 | 50416395 | 50416528 |
| chr19 | 50416609 | 50416723 |
| chr19 | 50417044 | 50417097 |
| chr19 | 50417171 | 50417269 |
| chr19 | 50417841 | 50417947 |
| chr2 | 29193223 | 29193922 |
| chr2 | 29196769 | 29196860 |
| chr2 | 29197541 | 29197676 |
| chr2 | 29207170 | 29207272 |
| chr2 | 29209785 | 29209878 |
| chr2 | 29213983 | 29214081 |
| chr2 | 29220705 | 29220835 |
| chr2 | 29222343 | 29222408 |
| chr2 | 29222516 | 29226234 |
| chr2 | 29226474 | 29227672 |
| chr2 | 29228883 | 29229066 |
| chr2 | 29232303 | 29232448 |
| chr2 | 29233564 | 29233696 |
| chr2 | 29239679 | 29239830 |
| chr2 | 29251104 | 29251267 |
| chr2 | 29275098 | 29275227 |
| chr2 | 29275401 | 29275496 |
| chr2 | 29296887 | 29297057 |
| chr2 | 29318303 | 29318404 |
| chr2 | 29320750 | 29320882 |
| chr2 | 29328349 | 29328481 |
| chr2 | 29383731 | 29383859 |
| chr2 | 29531914 | 29532116 |
| chr2 | 29694849 | 29695014 |
| chr2 | 29717577 | 29717697 |
| chr2 | 29919992 | 29920659 |
| chr2 | 47369505 | 47369581 |
| chr2 | 47373462 | 47373570 |
| chr2 | 47373807 | 47374048 |
| chr2 | 47375233 | 47375299 |
| chr2 | 47377013 | 47377077 |
| chr2 | 47378952 | 47379054 |
| chr2 | 47379768 | 47379969 |
| chr2 | 47385165 | 47385210 |
| chr2 | 47386571 | 47386613 |
| chr2 | 47403191 | 47403402 |
| chr2 | 47408400 | 47408555 |
| chr2 | 47410093 | 47410372 |
| chr2 | 47412413 | 47412560 |
| chr2 | 47414268 | 47414418 |
| chr2 | 47416295 | 47416429 |
| chr2 | 47429741 | 47429941 |
| chr2 | 47445547 | 47445657 |
| chr2 | 47463030 | 47463154 |
| chr2 | 47466657 | 47466808 |
| chr2 | 47470964 | 47471062 |
| chr2 | 47475024 | 47475270 |
| chr2 | 47476366 | 47476571 |
| chr2 | 47478271 | 47478519 |
| chr2 | 47480695 | 47480871 |
| chr2 | 47482778 | 47482949 |
| chr2 | 47783233 | 47783493 |
| chr2 | 47790926 | 47791123 |
| chr2 | 47795893 | 47796063 |
| chr2 | 47798610 | 47801155 |
| chr2 | 47803419 | 47803685 |
| chr2 | 47804909 | 47805027 |
| chr2 | 47805617 | 47805707 |
| chr2 | 47806203 | 47806358 |
| chr2 | 47806451 | 47806651 |
| chr2 | 47806778 | 47806860 |
| chr2 | 58159764 | 58159800 |
| chr2 | 58160107 | 58160179 |
| chr2 | 58161521 | 58161638 |
| chr2 | 58162865 | 58162947 |
| chr2 | 58163028 | 58163074 |
| chr2 | 58163433 | 58163517 |
| chr2 | 58165723 | 58165874 |
| chr2 | 58198578 | 58198662 |
| chr2 | 58204129 | 58204226 |
| chr2 | 58221941 | 58222042 |
| chr2 | 58226727 | 58226784 |
| chr2 | 58229813 | 58229874 |
| chr2 | 58232053 | 58232112 |
| chr2 | 58241217 | 58241313 |
| chr2 | 177230784 | 177232008 |
| chr2 | 177232391 | 177232583 |
| chr2 | 177233249 | 177233339 |
| chr2 | 177234004 | 177234271 |
| chr2 | 177264531 | 177264576 |
| chr2 | 208239841 | 208240023 |
| chr2 | 208243405 | 208243624 |
| chr2 | 208248347 | 208248680 |
| chr2 | 208251408 | 208251571 |
| chr2 | 214728675 | 214729008 |
| chr2 | 214730410 | 214730508 |
| chr2 | 214745066 | 214745159 |
| chr2 | 214745721 | 214745854 |
| chr2 | 214752446 | 214752555 |
| chr2 | 214767481 | 214767654 |
| chr2 | 214769231 | 214769312 |
| chr2 | 214780559 | 214781509 |
| chr2 | 214792296 | 214792445 |
| chr2 | 214797060 | 214797117 |
| chr2 | 214809411 | 214809569 |
| chr20 | 58840106 | 58840844 |
| chr20 | 58853265 | 58855333 |
| chr20 | 58891726 | 58891865 |
| chr20 | 58895611 | 58895684 |
| chr20 | 58898940 | 58898985 |
| chr20 | 58899948 | 58899955 |
| chr20 | 58903527 | 58903585 |
| chr20 | 58903671 | 58903791 |
| chr20 | 58905382 | 58905480 |
| chr20 | 58909161 | 58909216 |
| chr20 | 58909349 | 58909423 |
| chr20 | 58909520 | 58909579 |
| chr20 | 58909683 | 58909804 |
| chr20 | 58909950 | 58910081 |
| chr20 | 58910333 | 58910401 |
| chr20 | 58910682 | 58910829 |
| chr22 | 28687896 | 28687986 |
| chr22 | 28689134 | 28689215 |
| chr22 | 28694031 | 28694117 |
| chr22 | 28695126 | 28695242 |
| chr22 | 28695709 | 28695873 |
| chr22 | 28696900 | 28696987 |
| chr22 | 28699837 | 28699937 |
| chr22 | 28703434 | 28703636 |
| chr22 | 28709925 | 28710139 |
| chr22 | 28711908 | 28712017 |
| chr22 | 28719394 | 28719485 |
| chr22 | 28724976 | 28725124 |
| chr22 | 28725242 | 28725367 |
| chr22 | 28730419 | 28730548 |
| chr22 | 28734402 | 28734721 |
| chr3 | 12584492 | 12584677 |
| chr3 | 12604114 | 12604309 |
| chr3 | 36993547 | 36993663 |
| chr3 | 36996618 | 36996709 |
| chr3 | 37000954 | 37001053 |
| chr3 | 37004400 | 37004474 |
| chr3 | 37006990 | 37007063 |
| chr3 | 37008813 | 37008905 |
| chr3 | 37011819 | 37011862 |
| chr3 | 37012010 | 37012099 |
| chr3 | 37014431 | 37014544 |
| chr3 | 37017505 | 37017599 |
| chr3 | 37020309 | 37020463 |
| chr3 | 37025636 | 37026007 |
| chr3 | 37028783 | 37028932 |
| chr3 | 37040185 | 37040294 |
| chr3 | 37042267 | 37042331 |
| chr3 | 37047518 | 37047683 |
| chr3 | 37048516 | 37048609 |
| chr3 | 37048903 | 37049017 |
| chr3 | 37050485 | 37050653 |
| chr3 | 41224068 | 41224081 |
| chr3 | 41224525 | 41224753 |
| chr3 | 41224953 | 41225207 |
| chr3 | 41225333 | 41225572 |
| chr3 | 41225659 | 41225861 |
| chr3 | 41227207 | 41227352 |
| chr3 | 41233340 | 41233444 |
| chr3 | 41233528 | 41233867 |
| chr3 | 41234138 | 41234297 |
| chr3 | 41235723 | 41235843 |
| chr3 | 41236348 | 41236499 |
| chr3 | 41236587 | 41236709 |
| chr3 | 41238015 | 41238076 |
| chr3 | 41239133 | 41239342 |
| chr3 | 179198825 | 179199177 |
| chr3 | 179199689 | 179199899 |
| chr3 | 179201289 | 179201540 |
| chr3 | 179203543 | 179203789 |
| chr3 | 179204502 | 179204588 |
| chr3 | 179209594 | 179209700 |
| chr3 | 179210185 | 179210338 |
| chr3 | 179210430 | 179210565 |
| chr3 | 179219195 | 179219277 |
| chr3 | 179219570 | 179219735 |
| chr3 | 179219948 | 179220052 |
| chr3 | 179220985 | 179221157 |
| chr3 | 179224080 | 179224187 |
| chr3 | 179224699 | 179224821 |
| chr3 | 179225961 | 179226040 |
| chr3 | 179229271 | 179229442 |
| chr3 | 179230003 | 179230121 |
| chr3 | 179230224 | 179230376 |
| chr3 | 179234093 | 179234364 |
| chr4 | 1793934 | 1794043 |
| chr4 | 1799253 | 1799523 |
| chr4 | 1799746 | 1799812 |
| chr4 | 1801366 | 1801536 |
| chr4 | 1801619 | 1801743 |
| chr4 | 1801834 | 1802025 |
| chr4 | 1802913 | 1803064 |
| chr4 | 1803691 | 1803836 |
| chr4 | 1804329 | 1804520 |
| chr4 | 1804820 | 1804969 |
| chr4 | 1805354 | 1805476 |
| chr4 | 1805558 | 1805669 |
| chr4 | 1805749 | 1805940 |
| chr4 | 1806050 | 1806173 |
| chr4 | 1806256 | 1806327 |
| chr4 | 1806545 | 1806683 |
| chr4 | 1806828 | 1806934 |
| chr4 | 1807115 | 1807288 |
| chr4 | 54261073 | 54261432 |
| chr4 | 54263645 | 54263947 |
| chr4 | 54264897 | 54265069 |
| chr4 | 54267267 | 54267480 |
| chr4 | 54267530 | 54267761 |
| chr4 | 54273515 | 54273750 |
| chr4 | 54274819 | 54274993 |
| chr4 | 54277366 | 54277512 |
| chr4 | 54277874 | 54278026 |
| chr4 | 54278340 | 54278535 |
| chr4 | 54285819 | 54285983 |
| chr4 | 54295103 | 54295292 |
| chr4 | 54658014 | 54658081 |
| chr4 | 54695511 | 54695781 |
| chr4 | 54698283 | 54698565 |
| chr4 | 54699629 | 54699766 |
| chr4 | 54703723 | 54703892 |
| chr4 | 54707097 | 54707287 |
| chr4 | 54709423 | 54709539 |
| chr4 | 54723583 | 54723698 |
| chr4 | 54725856 | 54726050 |
| chr4 | 54727217 | 54727927 |
| chr4 | 54728010 | 54728121 |
| chr4 | 54729334 | 54729485 |
| chr4 | 54731327 | 54731419 |
| chr4 | 54731870 | 54731998 |
| chr4 | 54733069 | 54733192 |
| chr4 | 54736497 | 54736609 |
| chr4 | 54736720 | 54736820 |
| chr4 | 54737174 | 54737280 |
| chr4 | 54738428 | 54738557 |
| chr4 | 152322880 | 152323149 |
| chr4 | 152324183 | 152324394 |
| chr4 | 152326005 | 152326231 |
| chr4 | 152328207 | 152328389 |
| chr4 | 152329671 | 152329785 |
| chr4 | 152330731 | 152330868 |
| chr4 | 152332595 | 152332719 |
| chr4 | 152337801 | 152337936 |
| chr4 | 152346929 | 152347071 |
| chr4 | 152350041 | 152350124 |
| chr4 | 152352469 | 152352730 |
| chr4 | 152382188 | 152382335 |
| chr4 | 152411272 | 152411803 |
| chr5 | 1253727 | 1253831 |
| chr5 | 1254367 | 1254505 |
| chr5 | 1255286 | 1255411 |
| chr5 | 1258597 | 1258659 |
| chr5 | 1260473 | 1260600 |
| chr5 | 1264403 | 1264592 |
| chr5 | 1266463 | 1266535 |
| chr5 | 1268519 | 1268633 |
| chr5 | 1271118 | 1271204 |
| chr5 | 1272184 | 1272280 |
| chr5 | 1278640 | 1278796 |
| chr5 | 1279290 | 1279470 |
| chr5 | 1280157 | 1280338 |
| chr5 | 1282428 | 1282624 |
| chr5 | 1293312 | 1294666 |
| chr5 | 1294770 | 1296068 |
| chr5 | 112754890 | 112755025 |
| chr5 | 112766325 | 112766410 |
| chr5 | 112767188 | 112767390 |
| chr5 | 112775628 | 112775737 |
| chr5 | 112780789 | 112780903 |
| chr5 | 112792445 | 112792529 |
| chr5 | 112801278 | 112801383 |
| chr5 | 112815494 | 112815593 |
| chr5 | 112818965 | 112819344 |
| chr5 | 112821895 | 112821991 |
| chr5 | 112827107 | 112827247 |
| chr5 | 112827928 | 112828006 |
| chr5 | 112828855 | 112828972 |
| chr5 | 112834950 | 112835165 |
| chr5 | 112837552 | 112844126 |
| chr6 | 117288491 | 117288802 |
| chr6 | 117300973 | 117301137 |
| chr6 | 117308793 | 117308928 |
| chr6 | 117310080 | 117310281 |
| chr6 | 117311019 | 117311117 |
| chr6 | 117317142 | 117317272 |
| chr6 | 117318187 | 117318252 |
| chr6 | 117319867 | 117331483 |
| chr6 | 117331843 | 117332023 |
| chr6 | 117332983 | 117333163 |
| chr6 | 117334363 | 117334543 |
| chr6 | 117335143 | 117335263 |
| chr6 | 117335923 | 117336223 |
| chr6 | 117336463 | 117337340 |
| chr6 | 117341134 | 117341311 |
| chr6 | 117341399 | 117341632 |
| chr6 | 117342399 | 117342544 |
| chr6 | 117344059 | 117344262 |
| chr6 | 117352989 | 117353169 |
| chr6 | 117356628 | 117356915 |
| chr6 | 117357803 | 117358009 |
| chr6 | 117359808 | 117360011 |
| chr6 | 117360341 | 117360405 |
| chr6 | 117362602 | 117362865 |
| chr6 | 117365059 | 117365204 |
| chr6 | 117365580 | 117365741 |
| chr6 | 117366075 | 117366290 |
| chr6 | 117379058 | 117379159 |
| chr6 | 117383316 | 117383508 |
| chr6 | 117385682 | 117385861 |
| chr6 | 117386888 | 117386999 |
| chr6 | 117387779 | 117388034 |
| chr6 | 117389349 | 117389846 |
| chr6 | 117393223 | 117393321 |
| chr6 | 117394161 | 117394346 |
| chr6 | 117394615 | 117394738 |
| chr6 | 117396187 | 117396264 |
| chr6 | 117396914 | 117397116 |
| chr6 | 117403138 | 117403277 |
| chr6 | 117404279 | 117404428 |
| chr6 | 117409581 | 117409642 |
| chr6 | 117416257 | 117416317 |
| chr6 | 117418461 | 117418506 |
| chr6 | 117425533 | 117425656 |
| chr6 | 151807891 | 151808384 |
| chr6 | 151880633 | 151880791 |
| chr6 | 151944151 | 151944528 |
| chr6 | 152011634 | 152011814 |
| chr6 | 152060969 | 152061144 |
| chr6 | 152094363 | 152094588 |
| chr6 | 152098710 | 152098986 |
| chr7 | 5973398 | 5973542 |
| chr7 | 5977587 | 5977757 |
| chr7 | 5978595 | 5978696 |
| chr7 | 5982823 | 5982991 |
| chr7 | 5986758 | 5987620 |
| chr7 | 5989799 | 5989955 |
| chr7 | 5991972 | 5992057 |
| chr7 | 5995533 | 5995633 |
| chr7 | 5997325 | 5997423 |
| chr7 | 5999107 | 5999275 |
| chr7 | 6002452 | 6002636 |
| chr7 | 6003689 | 6003792 |
| chr7 | 6003971 | 6004058 |
| chr7 | 6005891 | 6006031 |
| chr7 | 6008996 | 6009019 |
| chr7 | 55019277 | 55019365 |
| chr7 | 55142285 | 55142437 |
| chr7 | 55143304 | 55143488 |
| chr7 | 55146605 | 55146740 |
| chr7 | 55151293 | 55151362 |
| chr7 | 55152545 | 55152664 |
| chr7 | 55154010 | 55154152 |
| chr7 | 55155829 | 55155946 |
| chr7 | 55156532 | 55156659 |
| chr7 | 55156758 | 55156843 |
| chr7 | 55157662 | 55157753 |
| chr7 | 55160138 | 55160338 |
| chr7 | 55161498 | 55161631 |
| chr7 | 55163732 | 55163823 |
| chr7 | 55165279 | 55165437 |
| chr7 | 55168522 | 55168529 |
| chr7 | 55170306 | 55170544 |
| chr7 | 55171174 | 55171213 |
| chr7 | 55172982 | 55173124 |
| chr7 | 55173920 | 55174043 |
| chr7 | 55192765 | 55192841 |
| chr7 | 55198716 | 55198863 |
| chr7 | 55200315 | 55200413 |
| chr7 | 55201187 | 55201355 |
| chr7 | 55201734 | 55201782 |
| chr7 | 55202516 | 55202765 |
| chr7 | 55205255 | 55205617 |
| chr7 | 116674320 | 116674381 |
| chr7 | 116678917 | 116678978 |
| chr7 | 116680954 | 116681015 |
| chr7 | 116687345 | 116687406 |
| chr7 | 116696862 | 116696923 |
| chr7 | 116699084 | 116700284 |
| chr7 | 116703253 | 116703314 |
| chr7 | 116714440 | 116714501 |
| chr7 | 116717126 | 116717187 |
| chr7 | 116730202 | 116730263 |
| chr7 | 116731667 | 116731859 |
| chr7 | 116739009 | 116739070 |
| chr7 | 116739949 | 116740084 |
| chr7 | 116740851 | 116741025 |
| chr7 | 116755354 | 116755515 |
| chr7 | 116757436 | 116757539 |
| chr7 | 116757637 | 116757774 |
| chr7 | 116758458 | 116758620 |
| chr7 | 116759336 | 116759490 |
| chr7 | 116763049 | 116763268 |
| chr7 | 116765302 | 116765363 |
| chr7 | 116769644 | 116769791 |
| chr7 | 116771497 | 116775111 |
| chr7 | 116775925 | 116775986 |
| chr7 | 116777388 | 116777469 |
| chr7 | 116778775 | 116778957 |
| chr7 | 116781987 | 116782097 |
| chr7 | 116783303 | 116783469 |
| chr7 | 116785408 | 116785469 |
| chr7 | 116790277 | 116790338 |
| chr7 | 116793442 | 116793503 |
| chr7 | 116795654 | 116795791 |
| chr7 | 116795886 | 116796124 |
| chr7 | 140726493 | 140726516 |
| chr7 | 140734596 | 140734770 |
| chr7 | 140739811 | 140739946 |
| chr7 | 140749286 | 140749418 |
| chr7 | 140754186 | 140754233 |
| chr7 | 140776911 | 140777088 |
| chr7 | 140777990 | 140778075 |
| chr7 | 140781575 | 140781693 |
| chr7 | 140783020 | 140783157 |
| chr7 | 140787547 | 140787584 |
| chr7 | 140794307 | 140794467 |
| chr7 | 140800361 | 140800481 |
| chr7 | 140801411 | 140801560 |
| chr7 | 140807959 | 140808062 |
| chr7 | 140808891 | 140808995 |
| chr7 | 140834608 | 140834872 |
| chr7 | 140850110 | 140850212 |
| chr7 | 140924565 | 140924703 |
| chr8 | 38413627 | 38413804 |
| chr8 | 38413917 | 38414023 |
| chr8 | 38414151 | 38414289 |
| chr8 | 38414558 | 38414629 |
| chr8 | 38414778 | 38414901 |
| chr8 | 38415869 | 38416060 |
| chr8 | 38417305 | 38417416 |
| chr8 | 38417869 | 38417991 |
| chr8 | 38418227 | 38418373 |
| chr8 | 38419532 | 38419735 |
| chr8 | 38421796 | 38421941 |
| chr8 | 38424508 | 38424699 |
| chr8 | 38426121 | 38426245 |
| chr8 | 38427920 | 38428093 |
| chr8 | 38428345 | 38428435 |
| chr8 | 38429681 | 38429948 |
| chr8 | 38440305 | 38440372 |
| chr8 | 38441870 | 38441931 |
| chr8 | 38457355 | 38457534 |
| chr8 | 38460788 | 38460849 |
| chr8 | 38461095 | 38461106 |
| chr8 | 38464797 | 38464858 |
| chr8 | 38467067 | 38467128 |
| chr9 | 5456113 | 5456165 |
| chr9 | 5457078 | 5457420 |
| chr9 | 5462833 | 5463121 |
| chr9 | 5465498 | 5465606 |
| chr9 | 5466769 | 5466829 |
| chr9 | 5467839 | 5467862 |
| chr9 | 21968228 | 21968242 |
| chr9 | 21968724 | 21968771 |
| chr9 | 21970901 | 21971208 |
| chr9 | 21974677 | 21974827 |
| chr9 | 21994138 | 21994331 |
| chr9 | 84670748 | 84670960 |
| chr9 | 84702158 | 84702233 |
| chr9 | 84702347 | 84702419 |
| chr9 | 84707843 | 84707912 |
| chr9 | 84710636 | 84710791 |
| chr9 | 84723572 | 84723709 |
| chr9 | 84724223 | 84724356 |
| chr9 | 84727653 | 84727959 |
| chr9 | 84741891 | 84741927 |
| chr9 | 84744972 | 84745073 |
| chr9 | 84751985 | 84752085 |
| chr9 | 84810541 | 84810579 |
| chr9 | 84861039 | 84861087 |
| chr9 | 84867242 | 84867431 |
| chr9 | 84871788 | 84871817 |
| chr9 | 84934161 | 84934292 |
| chr9 | 84948461 | 84948634 |
| chr9 | 84955282 | 84955517 |
| chr9 | 85020205 | 85020364 |
| chr9 | 85021251 | 85021437 |
| chr9 | 130862741 | 130863055 |
| chr9 | 130872107 | 130872233 |
| chr9 | 130872838 | 130873057 |
| chr9 | 130874846 | 130875072 |
| chr9 | 130883947 | 130885703 |
| chr9 | 136494432 | 136497578 |
| chr9 | 136498898 | 136498996 |
| chr9 | 136499090 | 136499279 |
| chr9 | 136500530 | 136500867 |
| chr9 | 136501747 | 136501913 |
| chr9 | 136502000 | 136502088 |
| chr9 | 136502250 | 136502508 |
| chr9 | 136503160 | 136503350 |
| chr9 | 136504651 | 136505124 |
| chr9 | 136505288 | 136505901 |
| chr9 | 136506526 | 136506639 |
| chr9 | 136506715 | 136506973 |
| chr9 | 136507304 | 136507437 |
| chr9 | 136507954 | 136508139 |
| chr9 | 136508231 | 136508385 |
| chr9 | 136508869 | 136509071 |
| chr9 | 136509732 | 136509961 |
| chr9 | 136510631 | 136510825 |
| chr9 | 136511151 | 136511271 |
| chr9 | 136513020 | 136513134 |
| chr9 | 136513370 | 136513557 |
| chr9 | 136514488 | 136514722 |
| chr9 | 136515268 | 136515420 |
| chr9 | 136515461 | 136515736 |
| chr9 | 136515959 | 136516114 |
| chr9 | 136517250 | 136517405 |
| chr9 | 136517730 | 136517957 |
| chr9 | 136518115 | 136518312 |
| chr9 | 136518569 | 136518844 |
| chr9 | 136519421 | 136519585 |
| chr9 | 136522828 | 136523208 |
| chr9 | 136523695 | 136523999 |
| chr9 | 136544023 | 136544102 |
| chr9 | 136545725 | 136546048 |
| chrX | 44873511 | 44873732 |
| chrX | 45020588 | 45020750 |
| chrX | 45034909 | 45035005 |
| chrX | 45059225 | 45059486 |
| chrX | 45063400 | 45063837 |
| chrX | 45069557 | 45070377 |
| chrX | 45076675 | 45076846 |
| chrX | 45079124 | 45079371 |
| chrX | 45083438 | 45083628 |
| chrX | 45090701 | 45090884 |
| chrX | 45110057 | 45110269 |
| chrX | 47562967 | 47563063 |
| chrX | 47563225 | 47563329 |
| chrX | 47564796 | 47564899 |
| chrX | 47564984 | 47565139 |
| chrX | 47565242 | 47565354 |
| chrX | 47566638 | 47566780 |
| chrX | 47566883 | 47566911 |
| chrX | 47566985 | 47567131 |
| chrX | 47567229 | 47567432 |
| chrX | 47568717 | 47568894 |
| chrX | 47568986 | 47569033 |
| chrX | 47569538 | 47569657 |
| chrX | 47569892 | 47570024 |
| chrX | 47570877 | 47571012 |
| chrX | 47571322 | 47571457 |
| chrX | 101353172 | 101353371 |
| chrX | 101356030 | 101356288 |
| chrX | 101374514 | 101374654 |

Probes highlighted in green indicate the overlapping target regions with the OPERA's panel.

**Table S2.** Oligonucleotide sequences used for sequencing library preparation.

| Oligonucleotide | Step | Sequence (5'-3') |
| --- | --- | --- |
| DL primer | Linear amplification | TCGTCGGCAGCGTCAGATGTGTATAAGAGACAG-Specific primer sequence-(DL-blocker) |
| Single strand adapter | ssDNA ligation | Phos-GNNTGNNTGNNTGNN (UMI) - CTGTCTCTTATACACATCTCCGAGCCCACGAGAC -index i7- ATCTCGTATGCCGTCTTCTGCTTG-Spacer C3 |
| i5 index primer | Indexing PCR | AATGATACGGCGACCACCGAGATCTACACAGGCTATA -index i5- TCGTCGGCAGCGTCAGATG |
| p7 primer (reverse primer) | Indexing PCR/dsDNA library PCR | CAAGCAGAAGACGGCATACGAGAT |
| p5 primer (forward primer) | dsDNA library PCR | AATGATACGGCGACCACCGAGATCTACAC |

**Table S3.** The specific primer sequences of each panel.

| Primer | Panel | Length (nt) | Specific primer sequence | Chr. | Start (HG38) | End (HG38) | Strand | Working Con.(nM) |
| --- | --- | --- | --- | --- | --- | --- | --- | --- |
| TP53_i07-3 | Plus | 26 | CTGTGTTATCTCCTAGGTTGGCTCTG | chr17 | 7674281 | 7674306 | Minus | 100 |
| TP53_i08-3 | Plus | 33 | CCTATCCTGAGTAGTGGTAATCTACTGGGACGG | chr17 | 7673819 | 7673851 | Minus | 50 |
| KRAS_i04-4 | Plus | 26 | GCTCAGGACTTAGCAAGAAGTTATGG | chr12 | 25225651 | 25225676 | Minus | 100 |
| KRAS_i02-2 | Plus/Mutation | 33 | GGCCTGCTGAAAATGACTGAATATAAACTTGTG | chr12 | 25245364 | 25245396 | Minus | 100 |
| HER2_i17-3 | Plus | 25 | CCCCAAGACCACGACCAGCAGAATG | chr17 | 39723352 | 39723376 | Minus | 50 |
| HER2_i21-4 | Plus | 27 | ACCAGCACGTTCCGAGCGGCCAAGTCC | chr17 | 39725087 | 39725113 | Minus | 50 |
| EGFR_i18-3 | Plus | 29 | TGCCAGGGACCTTACCTTATACACCGTGC | chr7 | 55174030 | 55174058 | Minus | 50 |
| EGFR_i19-3 | Plus/Mutation | 32 | CACATCGAGGATTTCCTTGTTGGCTTTCGGAG | chr7 | 55174792 | 55174823 | Minus | 60 |
| EGFR_i20-2 | Plus | 36 | GCCAATATTGTCTTTGTGTTCCCGGACATAGTCCAG | chr7 | 55181404 | 55181439 | Minus | 50 |
| EGFR_i20-3 | Plus | 23 | GTGAGGCAGATGCCCAGCAGGCG | chr7 | 55181335 | 55181357 | Minus | 50 |
| EGFR_i21-3 | Plus | 30 | CCTCCTTCTGCATGGTATTCTTTCTCTTCC | chr7 | 55191841 | 55191870 | Minus | 100 |
| TP53_i07-2 | Minus | 29 | AGTCTTCCAGTGTGATGATGGTGAGGATG | chr17 | 7674185 | 7674213 | Plus | 100 |
| TP53_i08-1 | Minus | 24 | GTGAGGCTCCCCTTTCTTGCGGAG | chr17 | 7673732 | 7673755 | Plus | 50 |
| KRAS_i04-2 | Minus | 40 | TTGCAGAAAACAGATCTGTATTTATTTCAGTGTTACTTAC | chr12 | 25225574 | 25225613 | Plus | 100 |
| KRAS_i02-1 | Minus | 36 | ATGGTCCTGCACCAGTAATATGCATATTAAAACAAG | chr12 | 25245232 | 25245267 | Plus | 100 |
| HER2_i17-1 | Minus | 25 | CCCACCCCAAACTAGCCCTCAATCC | chr17 | 39723271 | 39723295 | Plus | 80 |
| HER2_i20-1 | Minus | 24 | CCCTCTCAGCGTACCCTTGTCCCC | chr17 | 39724700 | 39724723 | Plus | 80 |
| HER2_i21-2 | Minus | 25 | AGGCCCTCCCAGAAGGTCTACATGG | chr17 | 39725008 | 39725032 | Plus | 80 |
| EGFR_i18-1 | Minus | 25 | ACACCCAGTGGAGAAGCTCCCAACC | chr7 | 55173936 | 55173960 | Plus | 100 |
| EGFR_i19-2 | Minus | 33 | CCAGTTAACGTCTTCCTTCTCTCTCTGTCATAG | chr7 | 55174689 | 55174721 | Plus | 100 |
| EGFR_i20-5 | Minus | 27 | CCTGCTGGGCATCTGCCTCACCTCCAC | chr7 | 55181337 | 55181363 | Plus | 50 |
| EGFR_i20-4 | Minus/Mutation | 27 | ATGCGAAGCCACACTGACGTGCCTCTC | chr7 | 55181255 | 55181281 | Plus | 50 |
| EGFR_i21-2 | Minus/ mutation | 30 | CCTGGCAGCCAGGAACGTACTGGTGAAAAC | chr7 | 55191760 | 55191789 | Plus | 50 |
| BRAF_i15-2 | Mutation | 29 | GGATCCAGACAACTGTTCAAACTGATGGG | chr7 | 140753287 | 140753315 | Plus | 100 |
| EGFR_i20-5 | Mutation | 28 | GCCTGCTGGGCATCTGCCTCACCTCCAC | chr7 | 55181336 | 55181363 | Plus | 40 |
| KRAS_i03-1 | Mutation | 27 | TCCCCAGTCCTCATGTACTGGTCCCTC | chr12 | 25227297 | 25227323 | Plus | 60 |
| NRAS_i03-1 | Mutation | 32 | CAGAGGAAGCCTTCGCCTGTCCTCATGTATTG | chr1 | 114713851 | 114713882 | Plus | 80 |
| PIK3CA_i10-1 | Mutation | 32 | AGAATCTCCATTTTAGCACTTACCTGTGACTC | chr3 | 179218326 | 179218357 | Minus | 70 |
| TP53_i05-1 | Mutation | 32 | GACTTTCAACTCTGTCTCCTTCCTCTTCCTAC | chr17 | 7675239 | 7675270 | Minus | 60 |
| TP53_i05-2 | Mutation | 25 | GCCATGGCCATCTACAAGCAGTCAC | chr17 | 7675113 | 7675137 | Minus | 100 |
| TP53_i06-1 | Mutation | 25 | CTGGCCCCTCCTCAGCATCTTATCC | chr17 | 7674945 | 7674969 | Minus | 50 |

**Table. S4**. Ligation efficiency and library conversion rate of unmodified and modified DL-primers.

| DL-Primer Sequence (5'-3') | 3' modification of Last Nucleotide with Specific base | base of Last Nucleotide | 3' modification of Last Nucleotide | Phosphorthioate | Input (copies) | Ligation efficiency | Library conversion rate |
| --- | --- | --- | --- | --- | --- | --- | --- |
| TCGTCGGCAGCGTCAGATGTGTATAAGAGACAGCCTGGCAGCCAGGAACGTACTGGTGAAAAC | OH | C | OH | 0 | 6000 | 1.2% | 0.12 |
| TCGTCGGCAGCGTCAGATGTGTATAAGAGACAGCCTGGCAGCCAGGAACGTACTGGTGAA*A*A*CT | T-spacer C3 | T | spacer C3 | 3 | 6000 | 30.0% | 2.67 |

* indicated the locations of phophorthioate modifications.

**Table. S5**. The performance of OPERA in detecting SNV and Indel in cell line samples.

| Template | VAF | Sensitivity | Mean VAF | BRAF V600E | PIK3CA E545K | NRAS Q61H | EGFR L858R | EGFR T790M | KRAS Q61H | KRAS G12C | TP53 S215G | ERBB2 A775-G776insYVMA | EGFR E746-A750del |
| --- | --- | --- | --- | --- | --- | --- | --- | --- | --- | --- | --- | --- | --- |
| Long | 0.0025% | 10% | 0.003% | 0.03% | 0 | 0 | 0% | 0 | 0 | 0 | 0% | 0 | 0 |
| Long | 0.0025% | 10% | 0.006% | 0 | 0 | 0.01% | 0% | 0.05% | 0 | 0 | 0% | 0 | 0 |
| Long | 0.0025% | 30% | 0.006% | 0.02% | 0 | 0.03% | 0.01% | 0 | 0 | 0 | 0% | 0 | 0 |
| Long | 0.0025% | 30% | 0.006% | 0.02% | 0 | 0 | 0.01% | 0.01% | 0 | 0 | 0.02% | 0 | 0 |
| Long | 0.0050% | 20% | 0.015% | 0 | 0.09% | 0% | 0% | 0.06% | 0 | 0 | 0% | 0 | 0 |
| Long | 0.0050% | 30% | 0.014% | 0.04% | 0 | 0.04% | 0.06% | 0 | 0 | 0 | 0% | 0 | 0 |
| Long | 0.0050% | 30% | 0.015% | 0.06% | 0 | 0% | 0% | 0.02% | 0 | 0.04% | 0 | 0.03% | 0 |
| Long | 0.0050% | 40% | 0.022% | 0 | 0 | 0 | 0 | 0.06% | 0 | 0.03% | 0.02% | 0.09% | 0.02% |
| Long | 0.0100% | 30% | 0.024% | 0.11% | 0.05% | 0 | 0.02% | 0 | 0.06% | 0 | 0 | 0 | 0 |
| Long | 0.0100% | 50% | 0.027% | 0.06% | 0.03% | 0.01% | 0.03% | 0.12% | 0 | 0 | 0.02% | 0 | 0 |
| Long | 0.0100% | 60% | 0.030% | 0.04% | 0.05% | 0.04% | 0.09% | 0.02% | 0.04% | 0 | 0 | 0.02% | 0 |
| Long | 0.0100% | 60% | 0.042% | 0.18% | 0.09% | 0.02% | 0.03% | 0.05% | 0 | 0 | 0% | 0.05% | 0 |
| Long | 0.0300% | 60% | 0.045% | 0 | 0.09% | 0.05% | 0.07% | 0.14% | 0 | 0 | 0.08% | 0 | 0.02% |
| Long | 0.0300% | 70% | 0.061% | 0.18% | 0 | 0.04% | 0.05% | 0.06% | 0 | 0 | 0.11% | 0.07% | 0.10% |
| Long | 0.0300% | 80% | 0.044% | 0.06% | 0.09% | 0% | 0.04% | 0.04% | 0 | 0.05% | 0.08% | 0.05% | 0.03% |
| Long | 0.0300% | 100% | 0.062% | 0.07% | 0.07% | 0.08% | 0.09% | 0.12% | 0.07% | 0.04% | 0.05% | 0.02% | 0.01% |
| Long | 0.1000% | 60% | 0.090% | 0.38% | 0.01% | 0.01% | 0.18% | 0.17% | 0 | 0.05% | 0.03% | 0.07% | 0 |
| Long | 0.1000% | 90% | 0.162% | 0.21% | 0.21% | 0.12% | 0.29% | 0.13% | 0.37% | 0.11% | 0% | 0.06% | 0.12% |
| Long | 0.1000% | 100% | 0.201% | 0.63% | 0.09% | 0.07% | 0.13% | 0.15% | 0.35% | 0.12% | 0.16% | 0.29% | 0.02% |
| Long | 0.1000% | 100% | 0.149% | 0.40% | 0.10% | 0.12% | 0.12% | 0.21% | 0.09% | 0.06% | 0.22% | 0.09% | 0.08% |
| Long | 0.3000% | 100% | 0.339% | 1.11% | 0.36% | 0.28% | 0.31% | 0.47% | 0.15% | 0.23% | 0.28% | 0.03% | 0.17% |
| Long | 0.3000% | 100% | 0.341% | 0.67% | 0.23% | 0.21% | 0.84% | 0.70% | 0.13% | 0.13% | 0.14% | 0.31% | 0.05% |
| Long | 0.3000% | 100% | 0.530% | 1.12% | 0.92% | 0.20% | 0.86% | 0.89% | 0.28% | 0.23% | 0.31% | 0.23% | 0.26% |
| Long | 0.3000% | 100% | 0.595% | 1.23% | 1.33% | 0.35% | 0.55% | 0.80% | 0.21% | 0.40% | 0.40% | 0.28% | 0.40% |
| Long | 1.0000% | 100% | 1.027% | 2.51% | 1.66% | 0.46% | 1.67% | 1.33% | 0.51% | 0.89% | 0.52% | 0.32% | 0.40% |
| Long | 1.0000% | 100% | 1.145% | 2.69% | 1.14% | 0.54% | 1.67% | 1.89% | 0.88% | 0.93% | 0.82% | 0.44% | 0.45% |
| Long | 1.0000% | 100% | 1.486% | 3.36% | 2.11% | 1.25% | 1.79% | 1.90% | 0.87% | 1.11% | 0.97% | 0.88% | 0.62% |
| Long | 1.0000% | 100% | 1.526% | 3.31% | 1.71% | 0.81% | 1.70% | 2.36% | 0.99% | 1.13% | 1.11% | 1.31% | 0.83% |
| NC | 0 | 0% | 0.000% | 0 | 0 | 0% | 0% | 0 | 0 | 0 | 0 | 0 | 0 |
| NC | 0 | 0% | 0.001% | 0 | 0 | 0% | 0% | 0.01% | 0 | 0 | 0% | 0 | 0 |
| NC | 0 | 0% | 0.002% | 0 | 0 | 0% | 0.01% | 0.01% | 0 | 0 | 0% | 0 | 0 |
| NC | 0 | 0% | 0.002% | 0 | 0 | 0 | 0% | 0.02% | 0 | 0 | 0% | 0 | 0 |
| short | 0.0025% | 10% | 0.004% | 0.04% | 0 | 0 | 0% | 0 | 0 | 0 | 0 | 0 | 0 |
| short | 0.0025% | 10% | 0.003% | 0 | 0 | 0% | 0.03% | 0 | 0 | 0 | 0 | 0 | 0 |
| short | 0.0025% | 10% | 0.007% | 0.05% | 0 | 0% | 0.02% | 0 | 0 | 0 | 0 | 0 | 0 |
| Short | 0.0025% | 10% | 0.003% | 0 | 0 | 0% | 0% | 0.03% | 0 | 0 | 0% | 0 | 0 |
| Short | 0.0050% | 10% | 0.009% | 0 | 0 | 0% | 0 | 0 | 0 | 0.09% | 0% | 0 | 0 |
| Short | 0.0050% | 20% | 0.010% | 0.04% | 0 | 0.06% | 0 | 0 | 0 | 0 | 0 | 0 | 0 |
| short | 0.0050% | 20% | 0.015% | 0 | 0.06% | 0% | 0 | 0.02% | 0.07% | 0 | 0 | 0 | 0 |
| Short | 0.0050% | 30% | 0.016% | 0.02% | 0.08% | 0.01% | 0% | 0.05% | 0 | 0 | 0% | 0 | 0 |
| short | 0.0100% | 20% | 0.017% | 0.08% | 0 | 0.02% | 0% | 0.03% | 0 | 0 | 0 | 0 | 0.04% |
| Short | 0.0100% | 30% | 0.011% | 0 | 0 | 0.01% | 0.02% | 0.03% | 0.05% | 0 | 0% | 0 | 0 |
| Short | 0.0100% | 40% | 0.037% | 0.08% | 0 | 0.05% | 0.09% | 0 | 0 | 0 | 0.15% | 0 | 0 |
| Short | 0.0100% | 40% | 0.025% | 0.05% | 0 | 0% | 0.09% | 0.05% | 0 | 0.06% | 0 | 0 | 0 |
| Short | 0.0300% | 50% | 0.037% | 0.15% | 0.08% | 0.02% | 0.04% | 0.03% | 0 | 0.03% | 0 | 0 | 0.02% |
| Short | 0.0300% | 60% | 0.042% | 0.06% | 0.11% | 0 | 0.10% | 0.06% | 0 | 0.06% | 0 | 0.03% | 0 |
| Short | 0.0300% | 70% | 0.049% | 0.16% | 0.09% | 0.01% | 0.06% | 0.05% | 0.05% | 0.03% | 0 | 0 | 0.04% |
| Short | 0.0300% | 70% | 0.050% | 0.05% | 0 | 0.05% | 0.21% | 0.09% | 0 | 0.05% | 0.03% | 0.02% | 0 |
| Short | 0.1000% | 70% | 0.088% | 0.21% | 0.28% | 0.09% | 0.03% | 0.17% | 0.06% | 0 | 0.02% | 0 | 0.02% |
| Short | 0.1000% | 70% | 0.060% | 0.07% | 0 | 0.13% | 0.12% | 0.13% | 0.09% | 0.02% | 0.01% | 0.03% | 0 |
| Short | 0.1000% | 80% | 0.131% | 0.30% | 0.17% | 0.09% | 0.22% | 0.06% | 0 | 0.32% | 0.03% | 0 | 0.12% |
| Short | 0.1000% | 80% | 0.136% | 0.31% | 0 | 0.24% | 0.17% | 0.23% | 0.14% | 0.13% | 0 | 0.07% | 0.07% |
| Short | 0.3000% | 80% | 0.172% | 0.58% | 0.20% | 0.26% | 0.22% | 0.25% | 0 | 0.10% | 0 | 0.08% | 0.03% |
| Short | 0.3000% | 90% | 0.279% | 0.52% | 0.58% | 0.32% | 0.33% | 0.26% | 0.26% | 0.14% | 0.21% | 0 | 0.17% |
| Short | 0.3000% | 100% | 0.439% | 1.15% | 0.41% | 0.55% | 0.77% | 0.58% | 0.07% | 0.42% | 0.20% | 0.20% | 0.04% |
| Short | 0.3000% | 100% | 0.502% | 0.96% | 0.73% | 0.33% | 1.00% | 0.74% | 0.37% | 0.41% | 0.12% | 0.12% | 0.24% |
| Short | 1.0000% | 100% | 1.023% | 1.94% | 2.26% | 0.83% | 1.38% | 1.05% | 0.52% | 1.13% | 0.74% | 0.24% | 0.14% |
| Short | 1.0000% | 100% | 1.568% | 3.52% | 2.46% | 1.09% | 1.92% | 2.55% | 1% | 1.61% | 0.60% | 0.31% | 0.62% |
| Short | 1.0000% | 100% | 1.353% | 3.32% | 1.85% | 0.99% | 2.10% | 1.80% | 0.83% | 1.47% | 0.45% | 0.24% | 0.48% |
| Short | 1.0000% | 100% | 1.488% | 3.03% | 2.35% | 1.10% | 2.26% | 1.89% | 1.12% | 1.38% | 0.81% | 0.48% | 0.46% |

NC, negative control; VAF, variant allelic frequency.

**Table. S6**. Theoretical detection ratio of OPERA.

| Mutation | Fragment length (bp) | Target length (bp) | Theoretical detection ratio (%) |
| --- | --- | --- | --- |
| L858R | 160 | 62 | 61.25 |
|  | 120 | 62 | 48.33 |
|  | 80 | 62 | 22.50 |
| T790M | 160 | 42 | 73.75 |
|  | 120 | 42 | 65.00 |
|  | 80 | 42 | 47.50 |

**Table S7**. Detailed patient information.

| Subject ID# | Gender | Age range | Disease | Subtype | Stage | T | N | M | L (cm) | W (cm) | H (cm) | Volume (cm^3^) | Vol-group (cm^3^) |
| --- | --- | --- | --- | --- | --- | --- | --- | --- | --- | --- | --- | --- | --- |
| P001 | M | ≤50 | NSCLC | ADC | III | 1b | 2 | 0 | 1.7 | 1.6 | 1.3 | 3.54 | 1-10 |
| P002 | F | 66-70 | NSCLC | ADC | III | 1c | 2 | 0 | 2.1 | 1.3 | 1.2 | 3.28 | 1-10 |
| P003 | F | 66-70 | NSCLC | ADC | I | 1c | 0 | 0 | 2.0 | 1.5 | 1.2 | 3.60 | 1-10 |
| P004 | M | 66-70 | NSCLC | ADC | III | 2a | 2 | 0 | 4.5 | 3.0 | 2.2 | 29.70 | >10 |
| P005 | M | 51-55 | NSCLC | ADC | I | 1c | 0 | 0 | 2.5 | 1.3 | 1.0 | 3.25 | 1-10 |
| P006 | M | 71-75 | NSCLC | ADC | I | 1c | 0 | 0 | 3.0 | 2.0 | 2.0 | 12.00 | >10 |
| P007 | M | 56-60 | NSCLC | ADC | I | 1b | 0 | 0 | 1.1 | 0.9 | 0.8 | 0.79 | <1 |
| P008 | F | 66-70 | NSCLC | ADC | I | 1c | 0 | 0 | 2.0 | 1.8 | 1.0 | 3.60 | 1-10 |
| P009 | F | 66-70 | NSCLC | ADC | I | 2a | 0 | 0 | 3.5 | 2.2 | 1.5 | 11.55 | >10 |
| P010 | M | 61-65 | NSCLC | ADC | II | 1b | 1 | 0 | 1.8 | 1.5 | 1.5 | 4.05 | 1-10 |
| P011 | F | 66-70 | NSCLC | ADC | I | 1c | 0 | 0 | 2.3 | 1.2 | 2.0 | 5.52 | 1-10 |
| P012 | M | 61-65 | NSCLC | ADC | I | 1b | 0 | 0 | 1.1 | 0.7 | 0.6 | 0.46 | <1 |
| P013 | F | 51-55 | NSCLC | ADC | I | 1b | 0 | 0 | 1.1 | 0.8 | 0.5 | 0.44 | <1 |
| P014 | M | 61-65 | NSCLC | ADC | III | 1c | 2 | 0 | 2.3 | 1.8 | 1.3 | 5.38 | 1-10 |
| P015 | F | 61-65 | NSCLC | ADC | I | 1c | 0 | 0 | 2.4 | 1.5 | 1.0 | 3.60 | 1-10 |
| P016 | F | 66-70 | NSCLC | ADC | I | 1c | 0 | 0 | 2.5 | 1.5 | 1.0 | 3.75 | 1-10 |
| P017 | M | 66-70 | NSCLC | ADC | I | 1c | 0 | 0 | 2.7 | 2.2 | 1.7 | 10.10 | >10 |
| P018 | F | 71-75 | NSCLC | ADC | I | 1b | 0 | 0 | 1.4 | 1.0 | 0.8 | 1.12 | 1-10 |
| P019 | M | 51-55 | NSCLC | ADC | III | 1c | 2 | 0 | 2.0 | 1.5 | 1.5 | 4.50 | 1-10 |
| P020 | M | ≤50 | NSCLC | ADC | I | 1b | 0 | 0 | 1.7 | 1.0 | 1.0 | 1.70 | 1-10 |
| P021 | F | 66-70 | NSCLC | ADC | I | 1b | 0 | 0 | 1.7 | 1.5 | 1.5 | 3.83 | 1-10 |
| P022 | M | 61-65 | NSCLC | ADC | I | 1c | 0 | 0 | 2.7 | 2.0 | 2.0 | 10.80 | >10 |
| P023 | F | 61-65 | NSCLC | ADC | I | 1b | 0 | 0 | 1.8 | 1.3 | 1.3 | 3.04 | 1-10 |
| P024 | M | 71-75 | NSCLC | SCC | I | 2b | 0 | 0 | 4.7 | 4.2 | 2.7 | 53.30 | >10 |
| P025 | F | 56-60 | NSCLC | ADC | I | 1b | 0 | 0 | 1.2 | 0.6 | 0.5 | 0.36 | <1 |
| P026 | F | 66-70 | NSCLC | ADC | I | 1b | 0 | 0 | 1.2 | 0.7 | 0.7 | 0.59 | <1 |
| P027 | F | 66-70 | NSCLC | ADC | I | 1b | 0 | 0 | 1.3 | 0.8 | 0.7 | 0.73 | <1 |
| P028 | F | 66-70 | NSCLC | ADC | I | 1b | 0 | 0 | 1.3 | 0.9 | 0.6 | 0.70 | <1 |
| P029 | F | 56-60 | NSCLC | ADC | I | 1b | 0 | 0 | 1.2 | 0.8 | 0.4 | 0.38 | <1 |
| P030 | F | ≤50 | NSCLC | ADC | I | 1a | 0 | 0 | 1.0 | 0.8 | 0.6 | 0.48 | <1 |
| P031 | M | 51-55 | NSCLC | ADC | I | 1b | 0 | 0 | 1.8 | 1.5 | 1.0 | 2.70 | 1-10 |
| P032 | M | 56-60 | NSCLC | ADC | II | 2a | 1 | 0 | 3.6 | 3.0 | 2.5 | 27.00 | >10 |
| P033 | M | 71-75 | NSCLC | SCC | I | 1b | 0 | 0 | 1.8 | 1.6 | 1.2 | 3.46 | 1-10 |
| P034 | F | 51-55 | NSCLC | ADC | I | 1b | 0 | 0 | 1.5 | 1.0 | 0.5 | 0.75 | <1 |
| P035 | F | 71-75 | NSCLC | ADC | I | 1b | 0 | 0 | 1.8 | 1.3 | 1.1 | 2.57 | 1-10 |
| P036 | M | ≤50 | Benign | Inflamed |  | / | / | / | / | / | / | / | / |
| P037 | M | 51-55 | NSCLC | ADC | III | 1c | 2 | 0 | 2.0 | 2.0 | 1.8 | 7.20 | 1-10 |
| P038 | M | 56-60 | NSCLC | ADC | III | 1c | 2 | 0 | 2.5 | 2.5 | 2.0 | 12.50 | >10 |
| P039 | F | 76-80 | NSCLC | ADC | I | 1b | 0 | 0 | 1.7 | 0.9 | 0.8 | 1.22 | 1-10 |
| P040 | F | ≤50 | NSCLC | ADC | I | 2a | 0 | 0 | 4.0 | 3.5 | 1.3 | 18.20 | >10 |

M, male; F, female; NSCLC, non-small cell lung cancer; ADC, adenocarcinoma; SCC, squamous cell carcinoma.

**Table S8.** Hot-spot depth of de-duplicated reads in 4 types samples.

| Sample type | Tumor tissue | Normal tissue | Blood cells | Plasma sample (including healthy group) |
| --- | --- | --- | --- | --- |
| *EGFR* 19del | 10706 | 7560 | 7144 | 4469 |
| *EGFR* 20ins | 6565 | 6178 | 4997 | 6084 |
| *EGFR* L858R | 11605 | 9013 | 7870 | 7410 |
| *EGFR* L861Q | 11037 | 8650 | 7483 | 7087 |
| *KRAS* G12D | 7135 | 6047 | 6265 | 4517 |
| *KRAS* G12R | 7157 | 6068 | 6281 | 4528 |
| *PIK3CA* E545D | 4073 | 3434 | 3196 | 2775 |
| *TP53* C135Y | 3539 | 3947 | 5532 | 5009 |
| *TP53* H179Y | 2724 | 2837 | 2598 | 2446 |
| *TP53* R175H | 2871 | 2966 | 2779 | 2604 |
| *TP53* S127T | 4134 | 4368 | 6360 | 5530 |
| ALL hot-spot Average | 6504 | 5552 | 5500 | 4769 |
| *EGFR* hot-spot Average | 9978 | 7850 | 6873 | 6263 |

**Table S9.** Determination of cutoff value in cfDNA, adjacent normal tissue and blood cell samples.

| Variant type | Substitution Type (plus strand) | Primer target strand | Primer strand Optimalization | Average single strand error rate (ASSER) | 10× ASSER | Mean VAF of Healthy cfDNA | Max VAF of Healthy cfDNA | VAF SD of Healthy cfDNA | Mean + 3×SD | Cutoff value |
| --- | --- | --- | --- | --- | --- | --- | --- | --- | --- | --- |
| *EGFR* 19del | deletion | Plus | / | / | / | 0.000% | 0.000% | 0.000% | 0.000% | 0.000% |
| *EGFR* 20ins | insertion | Minus | / | / | / | 0.000% | 0.000% | 0.000% | 0.000% | 0.000% |
| *EGFR* L858R | T>G | Minus | / | 4.08E-06 | 0.004% | 0.000% | 0.000% | 0.000% | 0.000% | 0.004% |
| *KRAS* G12D | C>T | Plus | No | 2.10E-04 | 0.210% | 0.023% | 0.120% | 0.039% | 0.141% | 0.210% |
| *KRAS* G12R | C>G | Plus | No | 8.63E-06 | 0.009% | 0.004% | 0.080% | 0.018% | 0.058% | 0.058% |
| *TP53* C135Y | C>T | Plus | No | 2.10E-04 | 0.210% | 0.009% | 0.070% | 0.021% | 0.073% | 0.210% |
| *TP53* R175H | C>T | Plus | No | 2.10E-04 | 0.210% | 0.305% | 1.140% | 0.322% | 1.270% | 1.270% |
| *EGFR* L861Q | T>A | Minus | Yes | 6.35E-07 | 0.001% | 0.000% | 0.000% | 0.000% | 0.000% | 0.001% |
| *PIK3CA* E545D | G>C | Plus | Yes | 3.64E-06 | 0.004% | 0.000% | 0.000% | 0.000% | 0.000% | 0.004% |
| *TP53* H179Y | G>A | Plus | Yes | 8.25E-05 | 0.082% | 0.000% | 0.000% | 0.000% | 0.000% | 0.082% |
| *TP53* S127T | A>T | Plus | Yes | 2.10E-06 | 0.002% | 0.000% | 0.000% | 0.000% | 0.000% | 0.002% |
